# Determinants of health-related quality of life in healthy children and adolescents during the COVID-19 pandemic: results from a prospective longitudinal cohort study

**DOI:** 10.1101/2023.08.25.23294563

**Authors:** Sarah R Haile, Gabriela P Peralta, Alessia Raineri, Sonja Rueegg, Agne Ulyte, Milo A Puhan, Thomas Radtke, Susi Kriemler

## Abstract

**Purpose:** Understanding health-related quality of life (HRQOL) in children and adolescents, during a pandemic and afterwards, aids in understanding how circumstances in their lives impact their well-being. We aimed to identify determinants of HRQOL from a broad range of biological, psychological and social factors in a large longitudinal population-based sample.

**Methods:** Data was taken from a longitudinal sample (n = 1843) of children and adolescents enrolled in the prospective school-based cohort study *Ciao Corona* in Switzerland. The primary outcome was HRQOL, assessed using the KINDL total score and its subscales (each from 0, worst, to 100, best). Potential determinants, including biological (physical activity, screen time, sleep, etc), psychological (sadness, anxiousness, stress) and social (nationality, parents’ education, etc) factors, were assessed in 2020 and 2021, and HRQOL in 2022. Determinants were identified in a data-driven manner using recursive partitioning to define homogeneous subgroups, stratified by school level.

**Results:** Median KINDL total score in the empirically identified subgroups ranged from 68 to 83 in primary school children and from 69 to 82 in adolescents in secondary school. The psychological factors sadness, anxiousness and stress in 2021 were identified as the most important determinants of HRQOL in both primary and secondary school children. Other factors, such as physical activity, screen time, chronic health conditions or nationality, were determinants only in individual subscales.

**Conclusion:** Recent mental health, more than biological, physical or social factors, played a key role in determining HRQOL in children and adolescents during pandemic times. Public health strategies to improve mental health may therefore be effective in improving HRQOL in this age group.

**What is known:**

- Assessing health-related quality of life (HRQOL) in children and adolescents aids in understanding how life circumstances impact their well-being.
- HRQOL is a complex construct, involving biological, psychological and social factors. Factors driving HRQOL in children and adolescents are not often studied in longitudinal population-based samples.

**What is new:**

- Mental health (stress, anxiousness, sadness) played a key role in determining HRQOL during the coronavirus pandemic, more than biological or social factors.
- Public health strategies to improve mental health may be effective in improving HRQOL in children.

## Introduction

Children and adolescents are formed by the environment they live in and their social engagement with those around them. The assessment of health-related quality of life (HRQOL) allows us to better understand how circumstances in their lives impact their well-being, especially during disasters like the coronavirus disease 2019 (COVID-19) pandemic. Many children and adolescents remain resilient over time or may recover rapidly. However, others may suffer from multiple stressors (e.g. illness, disruption of the family system, isolation, social separation from peers and home confinement) thereby impacting their short and long-term mental health and well-being [1]. Understanding HRQOL and building knowledge about physical, emotional and social challenges that children and adolescents may have experienced during the pandemic will allow to raise Public Health awareness and build the foundation for action now and in future difficult periods that may evolve.

Various studies have shown that HRQOL in children and adolescents has worsened during the COVID-19 pandemic [2–5], though there is some evidence that it has at least partially recovered to prepandemic levels [6]. HRQOL is a complex construct, defined as a subjective perception that an individual has about the impact their health has on their life, involving not only biological factors (e.g. body mass index (BMI), or chronic health conditions) [7], but also psychological (e.g. stress, anxiety or depression) and social factors (e.g. family support, social integration, or family atmosphere) [8,9]. In line with this bio-psycho-social construct [8], several studies have demonstrated positive associations between physical activity (PA) and HRQOL [10–12], and negative associations between BMI and HRQOL [13,14]. Other studies have explored associations with screen time [15–17], sleep [18], self-esteem and emotions [19], parents’ education and family wealth [20], and nationality [21]. While determinants of HRQOL in children with a range of chronic health conditions have been previously studied, generally prior to the pandemic, fewer have sought to identify possible determinants of HRQOL in healthy children, often with small sizes [12,15,21]. Surprisingly few studies took a global view on HRQOL in youth by trying to understand the influence of the broader bio-psycho-social construct on their well-being and teasing out which factors alone or in combination are most influential.

Using data from the Ciao Corona study, a prospective school-based cohort study of children and adolescents during the COVID-19 pandemic (2020 - 2022), we aimed to identify determinants of HRQOL at the end of the pandemic, June 2022. Using conditional inference trees, we aimed to identify both individual determinants and patterns of determinants that indicate clusters of children and adolescents with similar HRQOL in a large longitudinal population-based sample.

## Methods

### Study

The data for this analysis come from the school-based longitudinal cohort study Ciao Corona [22], in which 55 randomly selected schools (primary school grades 1-6 and secondary school grades 7-9, ages 6-17 years) in the canton of Zurich, the largest canton in Switzerland of approximately 1.5 million inhabitants (18% of the total Swiss population), took part. Subjects (or their parents) were also asked to fill out a baseline questionnaire at the time of their first antibody test, and to complete follow-up questionnaires on a periodic basis (July 2020, January 2021, March 2021, September 2021, and July 2022). The analysis set of this study included children and adolescents who had KINDL total scores in June 2022 and at least one questionnaire filled out earlier during the pandemic.

The study was approved by the ethical committee of the canton of Zurich (2020-01336), and the study design has been published elsewhere [22] (ClinicalTrials.gov identifier: NCT04448717). All participants provided written informed consent before being enrolled in the study. Results relating to lifestyle behaviors [23] and HRQOL [24] have been reported previously.

### Outcomes

The primary outcome was HRQOL in June 2022, assessed using the KINDL questionnaire, filled out either by primary school students and parents together, or by students in secondary school on their own. KINDL is a reliable and valid measure of HRQOL in children and adolescents [25,26], on a scale from 0 (worst) to 100 (best). It has 6 subscales, each from 0 to 100: physical, emotional, self-esteem, family, friends, and school. Several slightly different versions of KINDL are available, of which the parent version for children 7-17 years old was used [27]. Age group was determined by the highest grade level achieved during the study period. Children who were in 6th grade or lower were in the primary school group (having never gone to secondary school), while those who were in at least 7th grade by 2022 were in the secondary school group, even if they were still in primary school in 2020.

Possible determinants of HRQOL were pulled from questionnaires and categorized into three categories: biological, psychological and social determinants, as has previously been described as a model for HRQOL [7,8,28,29] (Figure 1). Biological variables included sex (male, female, other), BMI, PA, screen time (ST), sleep duration, presence of chronic health conditions, and symptoms possibly compatible with post-COVID-19 condition (also known as Long Covid). BMI was calculated according to weight and height, and compared with the standard Swiss population [30] to derive z-scores. BMI was then categorized as overweight if its z-score was 1 or higher. PA, ST and sleep were recorded in hours per week, which were then compared with World Health Organization recommendations [31] (*≥* 1 h/day of PA, *≤* 2 h/day of ST, and 9-11 h/night of sleep for 6-13 year olds or 8-10 h/night for 14-16 year olds). Chronic health conditions included asthma, celiac disease, neurodermatitis, type I diabetes, inflammatory bowel disease, hypertension, attention deficit hyperactivity disorder, epilepsy, joint disorders, and depression / anxiety. Possible post-COVID-19 condition [32,33] was identified if participants reported any number of symptoms lasting 3 months or longer that might be related to a COVID-19 infection in seropositive participants.

**Figure 1:**
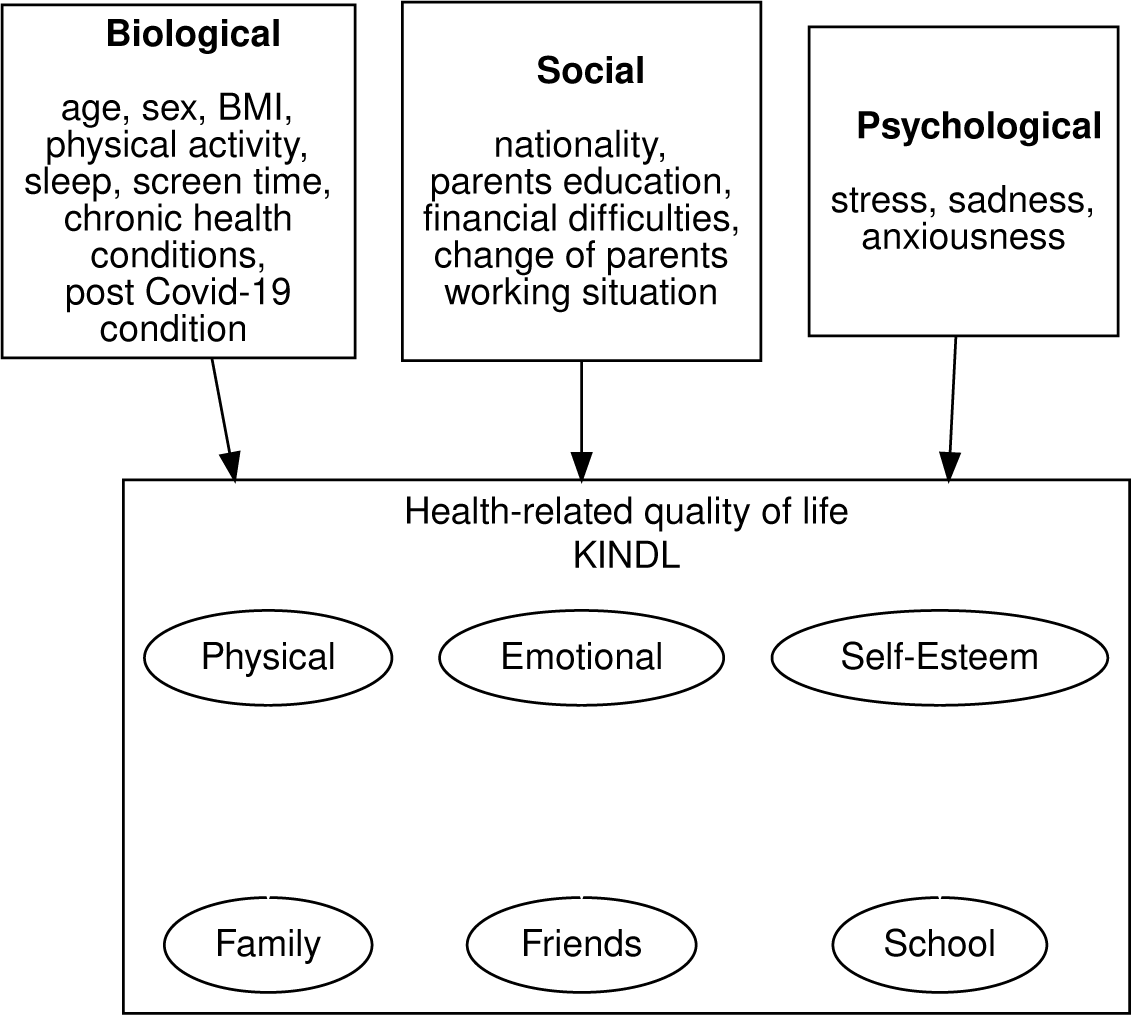
Potential determinants of health-related quality of life (HRQOL), grouped into biological, social and psychological factors.

Psychological variables included sadness, anxiousness and stress that were taken from Health-Behaviour in School-Aged Children questionnaires [34]. For sadness, children were asked how often they felt sad or depressed in the last 6 months (daily, multiple times per week, once per week, once per month, seldom or never). For anxiousness, they were asked how often in the last six months they felt scared or anxious (same responses as for depression). For stress, parents were asked how they would assess the level of stress in the child’s life on a scale from 1 (no stress) to 6 (extreme stress).

Social variables collected were parents’ nationality (at least 1 Swiss parent vs other), parents’ highest education level (at least one with college prep high school or university vs compulsory high school or professional school or lower), presence of household financial difficulties (yes/no), and change of parents’ working situation (reduction in or loss of work, vs no change), or change or loss of employment due to parents’ health [35].

Parents of participants were asked by email to fill out, with their child, a baseline questionnaire online at the time of their first serological test (June/July 2020, Oct/Nov 2020, Mar 2021, Nov/Dec 2021, June 2022). For the HRQOL, sadness, anxiousness, stress and physical activity questions, parents were specifically asked to answer them together with their child, Thereafter, they were invited to fill out an online follow-up questionnaire at semi-regular intervals (Sept/Oct 2020, Jan 2021, Mar 2021, Sept 2021, Dec 2021, June 2022). While the baseline questionnaires asked a number of demographic details (e.g. parents’ nationality and education levels), there was significant overlap in content between the baseline and follow-up questionnaires. For the KINDL questions, as well as those relating to sadness, anxiousness, and stress, . For data analysis, timepoints were grouped by year: 2019 (retrospective questions in the June 2020 questionnaire relating to the pre-pandemic period), 2020, 2021 and 2022. HRQOL was taken from the June 2022 questionnaire while possible determinants were taken from 2019-2021. If multiple questionnaires existed for a subject in the same year with the same question, the mean or most frequent response was taken. Further details on the questionnaires used in Ciao Corona are found in Online Resource 1.

### Statistical Methods

KINDL scores were summarized as median [Interquartile range (IQR)]. We used conditional inference trees [36,37] estimated by binary recursive partitioning to identify possible determinants of HRQOL in children and adolescents. This statistical method identifies subgroups where stratifying by possible predictor variable produces a statistically significant difference in the outcome variable. For further details, see Online Resource 2. This procedure was repeated for the KINDL total score as well as for each of its subscales, and stratified by age group (primary vs secondary school). As a sensitivity analysis, multiple imputation using chained equations was used to impute missing covariates [38] (m = 100), and then recursive partitioning was used to identify significant predictors of HRQOL in each of the imputed datasets. We then counted how often each variable was included in the model selection procedure, with more important variables appearing more often than variables which are not determinants. All analysis was performed in R (R version 4.3.2 (2023-10-31)) using the packages `partykit` [36,39] and `mice` [40].

## Results

There were 1843 children and adolescents who had KINDL total scores in June 2022 and at least one questionnaire filled out since June 2020 (Table 1). Approximately 8% of children were overweight, and 76% had highly educated parents. KINDL total scores remained stable in the period 2020 - 2022 (median primary school in 2020 82.3 [IQR 77.1 - 86.5], in 2022 80.2 [74.0 - 85.4], and in secondary school 2020 79.2 [72.9 - 85.1] and 2020 74.0 [67.7 - 81.2]), but were somewhat lower in secondary school children than in those remaining in primary school (Online Resource 2: Figure S1). Covariates considered to be potential determinants of HRQOL are displayed graphically in Figure 1 and listed in Table 1 by age group and timepoint (2019, 2020, or 2021).

**Table 1:** Covariates considered as potential determinants of health-related quality of life (HRQOL), by age group and timepoint. Sadness and Anxiousness were assessed on a scale from 1 (daily) to 5 (rarely or never). Stress was assessed on a scale from 1 (no stress) to 6 (extreme stress). ’na’ indicates not assessed.

| Characteristic | primary school |  |  |  | secondary school |  |  |  |
| --- | --- | --- | --- | --- | --- | --- | --- | --- |
|  | baseline | 2019 | 2020 | 2021 | baseline | 2019 | 2020 | 2021 |
| **Biological** |  |  |  |  |  |  |  |  |
| Sex (male) | 49%<br>(441/891) |  |  |  | 49%<br>(466/952) |  |  |  |
| Chronic health conditions | 12%<br>(109/891) |  |  |  | 16%<br>(149/952) |  |  |  |
| Symptoms compatible with post Covid-19 condition |  | na | na | 1.0%<br>(9/891) |  | na | na | 2.2%<br>(21/952) |
| Overweight / Obese |  | na | 10%<br>(40/389) | 8.0%<br>(29/361) |  | na | 12%<br>(60/520) | 7.4%<br>(34/461) |
| Physical activity guidelines met |  | 82%<br>(320/390) | 63%<br>(245/388) | 66%<br>(435/661) |  | 80%<br>(416/517) | 54%<br>(277/510) | 55%<br>(395/723) |
| Screen time guidelines met |  | 98%<br>(382/389) | 85%<br>(334/393) | 95%<br>(626/657) |  | 87%<br>(450/518) | 51%<br>(266/517) | 54%<br>(388/721) |
| Sleep guidelines met |  | 93%<br>(364/393) | 90%<br>(354/394) | 89%<br>(586/658) |  | 68%<br>(354/517) | 80%<br>(415/516) | 50%<br>(359/722) |
| **Psychological** |  |  |  |  |  |  |  |  |
| Sadness |  | na | 4.1<br><U+00B1><br>1.0 (386) | 4.3<br><U+00B1><br>0.8 (659) |  | na | 4.3<br><U+00B1><br>0.9 (518) | 4.1<br><U+00B1><br>0.9 (722) |
| Anxiousness |  | na | 4.5<br><U+00B1><br>0.9 (383) | 4.5<br><U+00B1><br>0.8 (658) |  | na | 4.6<br><U+00B1><br>0.8 (519) | 4.4<br><U+00B1><br>0.8 (723) |
| Stress |  | na | 2.1<br><U+00B1><br>1.0 (363) | 2.2<br><U+00B1><br>1.0 (652) |  | na | 2.9<br><U+00B1><br>1.2 (480) | 2.8<br><U+00B1><br>1.1 (716) |
| **Social** |  |  |  |  |  |  |  |  |
| Swiss nationality | 85%<br>(739/869) |  |  |  | 88%<br>(805/918) |  |  |  |
| High parental education | 81%<br>(698/861) |  |  |  | 72%<br>(647/903) |  |  |  |
| Household financial difficulties |  | na | na | 1.7%<br>(15/891) |  | na | na | 2.1%<br>(20/952) |
| Change in parents' working situation for health reasons |  | 0%<br>(0/891) | na | 0.6%<br>(5/891) |  | 0%<br>(0/951) | na | 0.2%<br>(2/952) |
| Change in parents' working situation |  | 0%<br>(0/891) | 18%<br>(161/891) | 2.0%<br>(18/891) |  | 0%<br>(0/951) | 19%<br>(177/952) | 1.7%<br>(16/952) |

<sup>1</sup> % (n/N); Mean ± SD (N)

We first examined KINDL total score in primary school children, which ranged from 38 to 100 with an interquartile range (IQR) of 11.5 points. Most of the variation in KINDL total score in 2022 in primary school children was explained by stress, sadness, and anxiousness in 2021 (Figure 2). Median KINDL total score in the identified subgroups ranged from 66 [IQR 57 to 73] (in those with frequent sadness, frequent anxiousness and moderate to high stress) to 83 [78 to 89] (in those reporting no stress), a difference which corresponded to 1.5 times the overall IQR. When repeating the analysis on each of the KINDL subscales (physical, emotional, self-esteem, family, friends, school), the same variables were generally chosen, along with sex and chronic health conditions (Figure 3, see boxes denoted “P” or “P, S”). Notably, variables from 2021 were more often chosen than their 2020 counterparts.

**Figure 2:**
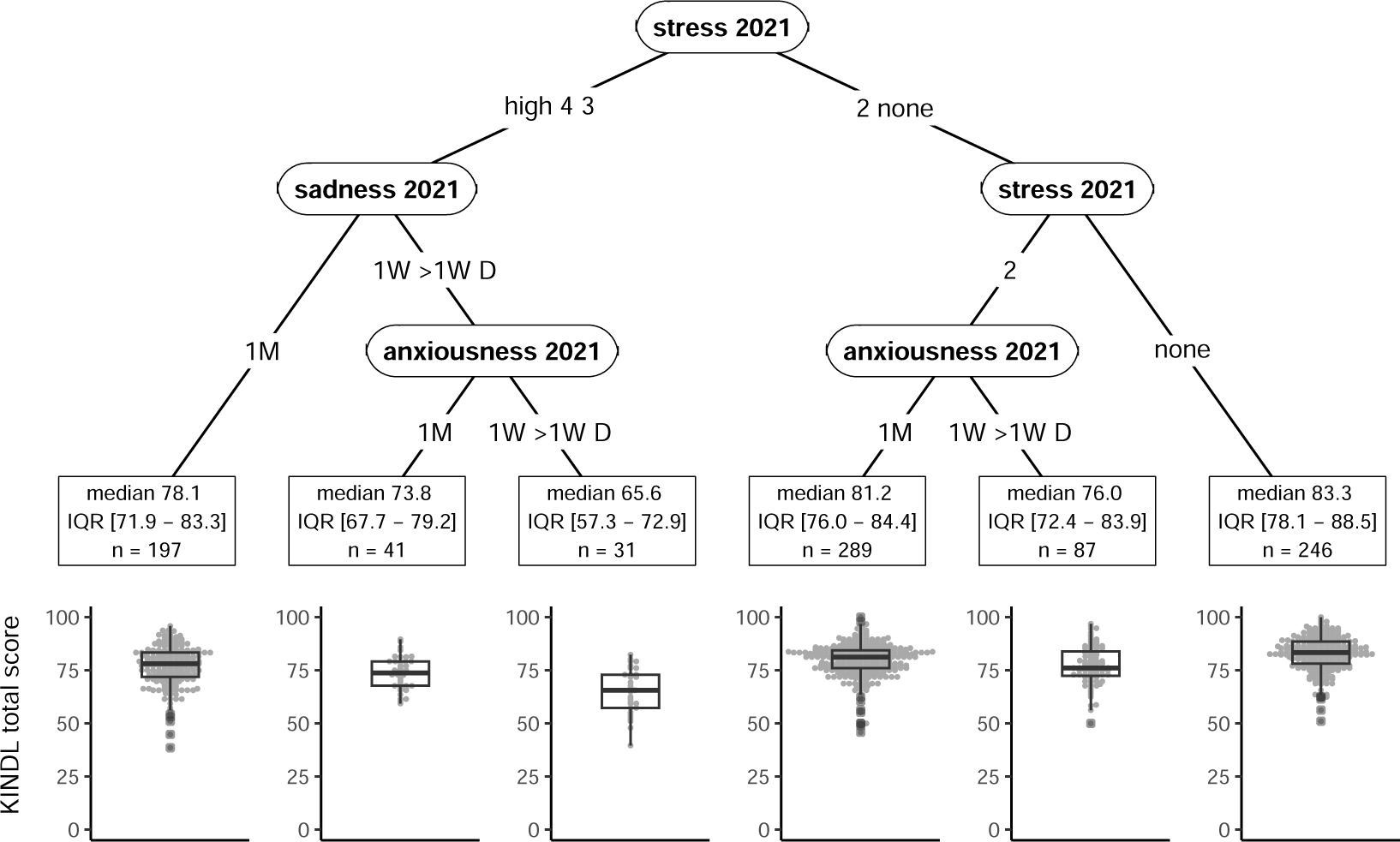
Recursive partitioning tree for health-related quality of life (HRQOL, KINDL total score) in 2022 in primary school children. Identified determinants are stress, sadness and anxiousness in 2021. Sadness and anxiousness could have occurred once per month (1M), once per week (1W), more than once per week (>1W) or daily (D). Stress was considered on a 5-point scale from 1 (no stress) to 4 (high stress). Other variables included in the model could not be used to create more homogeneous groups with respect to KINDL total score. For each subgroup, median KINDL total score, interquartile range (IQR), mean ± standard deviation and sample size (n) are given.

**Figure 3:**
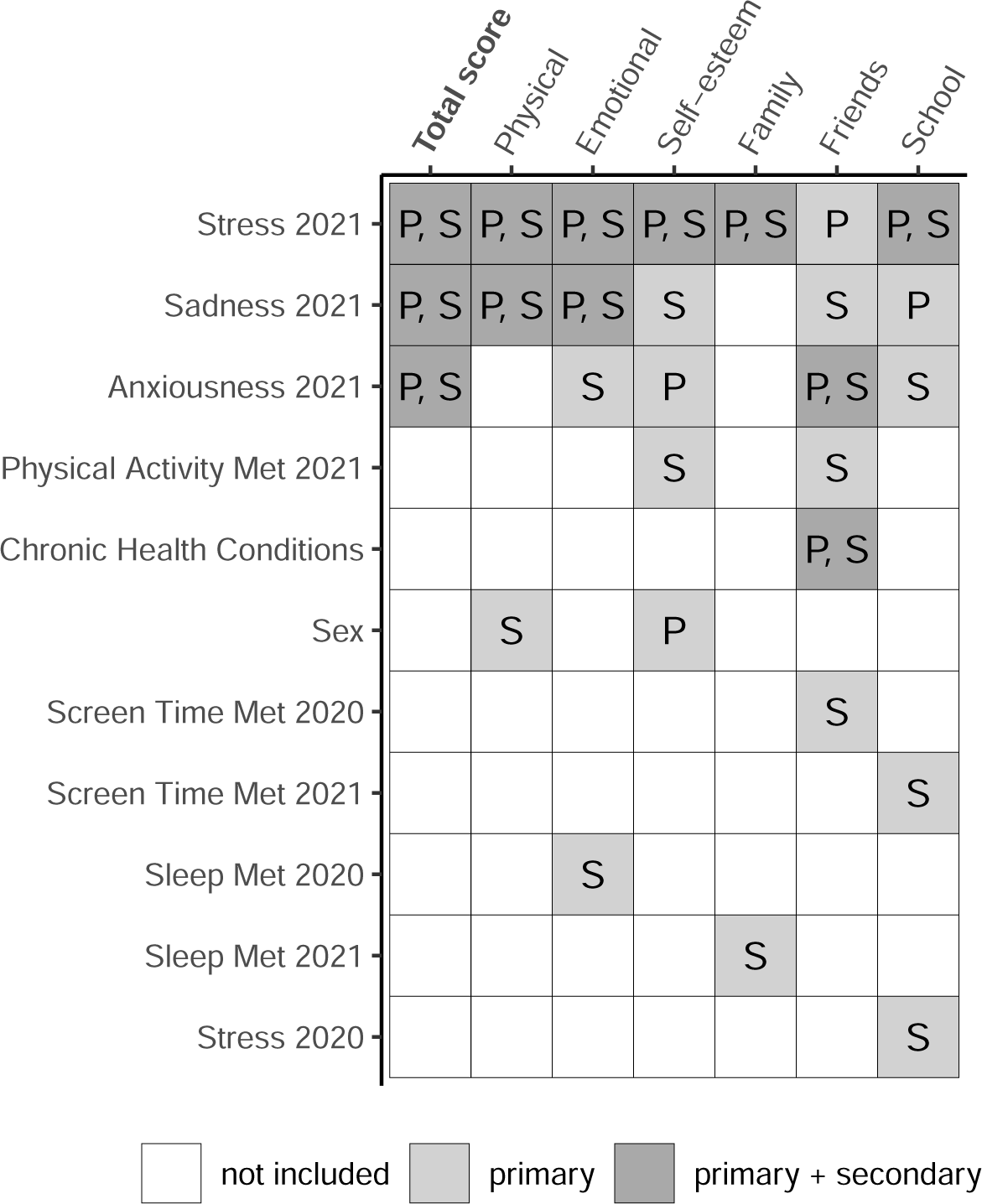
Overview of all variables identified as determinants of health-related quality of life assessed by the KINDL total score and its subscales: physical, emotional, self-esteem, family, friends, and school. ’P’ indicate variables identified for primary school only, ’S’ for secondary school only and ’P, S’ for both primary and secondary school. Variables not shown were not identified for any of the subscales.

Next we examined KINDL total score in secondary school children, ranging from 33 to 99 with IQR = 13.5, where most of the variation was explained by stress, anxiousness and sadness (Figure 4). Median KINDL total score in the identified subgroups ranged from 68 [IQR 62 to 73] (among those with moderate to high stress and frequent anxiety) to 80 [73 to 87] (among those with no stress and infrequent sadness), a difference of 0.9 IQR. Repeating the analysis for secondary school children on each of the subscales, sex, PA, sleep, ST, and chronic health conditions were additionally identified as predictors of various subscales (Figure 3, see boxes denoted “S” or “P, S”). However, none of these additional factors were identified as determinants for overall HRQOL in secondary school children.

**Figure 4:**
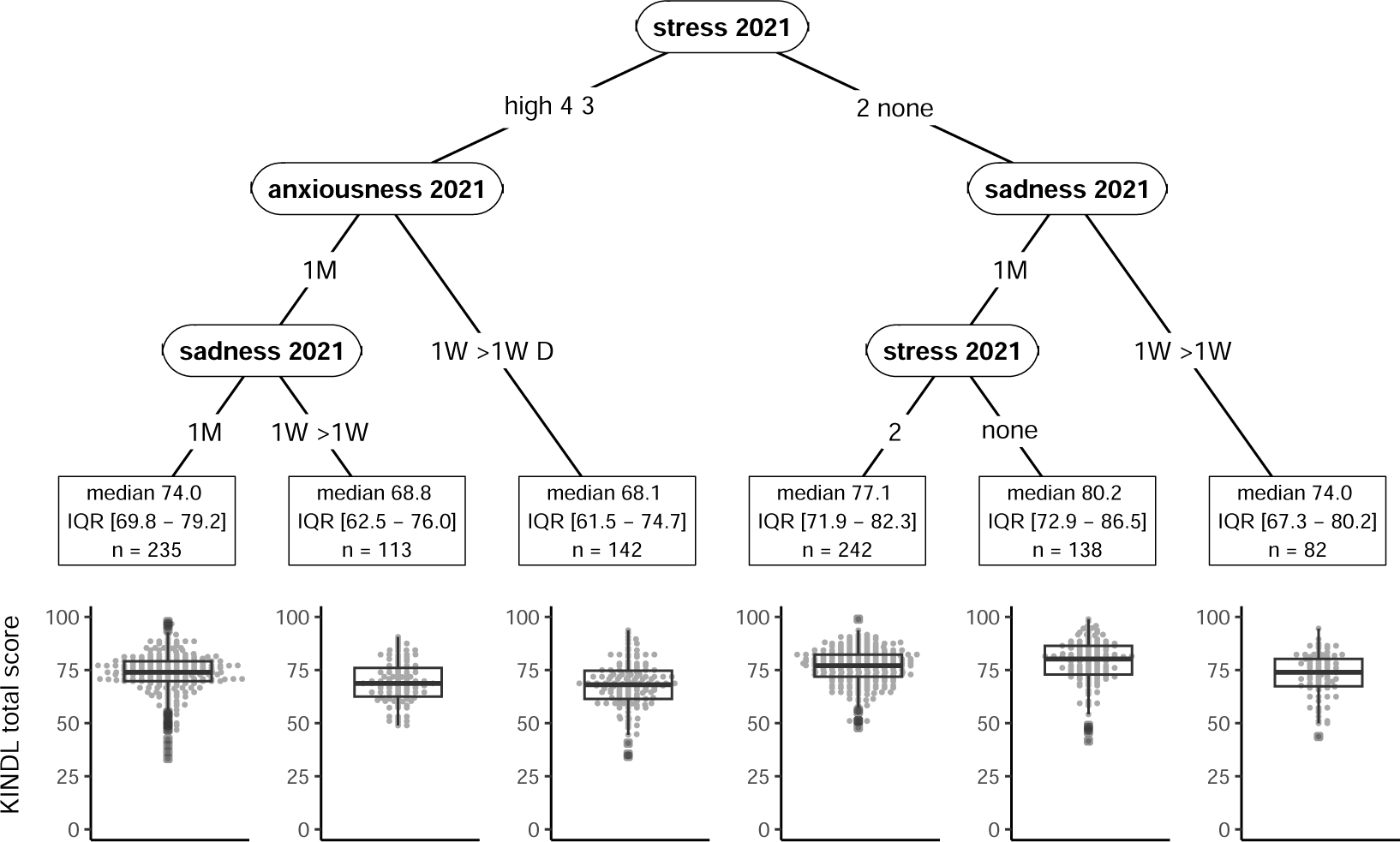
Recursive partitioning tree for health-related quality of life (HRQOL, KINDL total score) in 2022 in secondary school children. Identified determinants are stress, sadness, anxiousness, and self-rated health in 2021. Sadness and anxiousness could have occurred once per month (1M), once per week (1W), more than once per week (>1W) or daily (D). Stress was considered on a 5-point scale from 1 (no stress) to 4 (high stress). Self-rated health was rated as excellent, good, moderately good or bad. Other variables included in the model could not be used to create more homogeneous groups with respect to KINDL total score. For each subgroup, median KINDL total score, interquartile range (IQR), mean ± standard deviation and sample size (n) are given.

As there is some uncertainty in recursive partitioning where models may not always choose the same factors in the presence of missing data, we repeated the model for total KINDL score 100 times each for primary and secondary school children (Figure 5, Online Resource 2: Tables S4 and S5). For primary school children, the models most often included stress, anxiousness and sadness.

**Figure 5:**
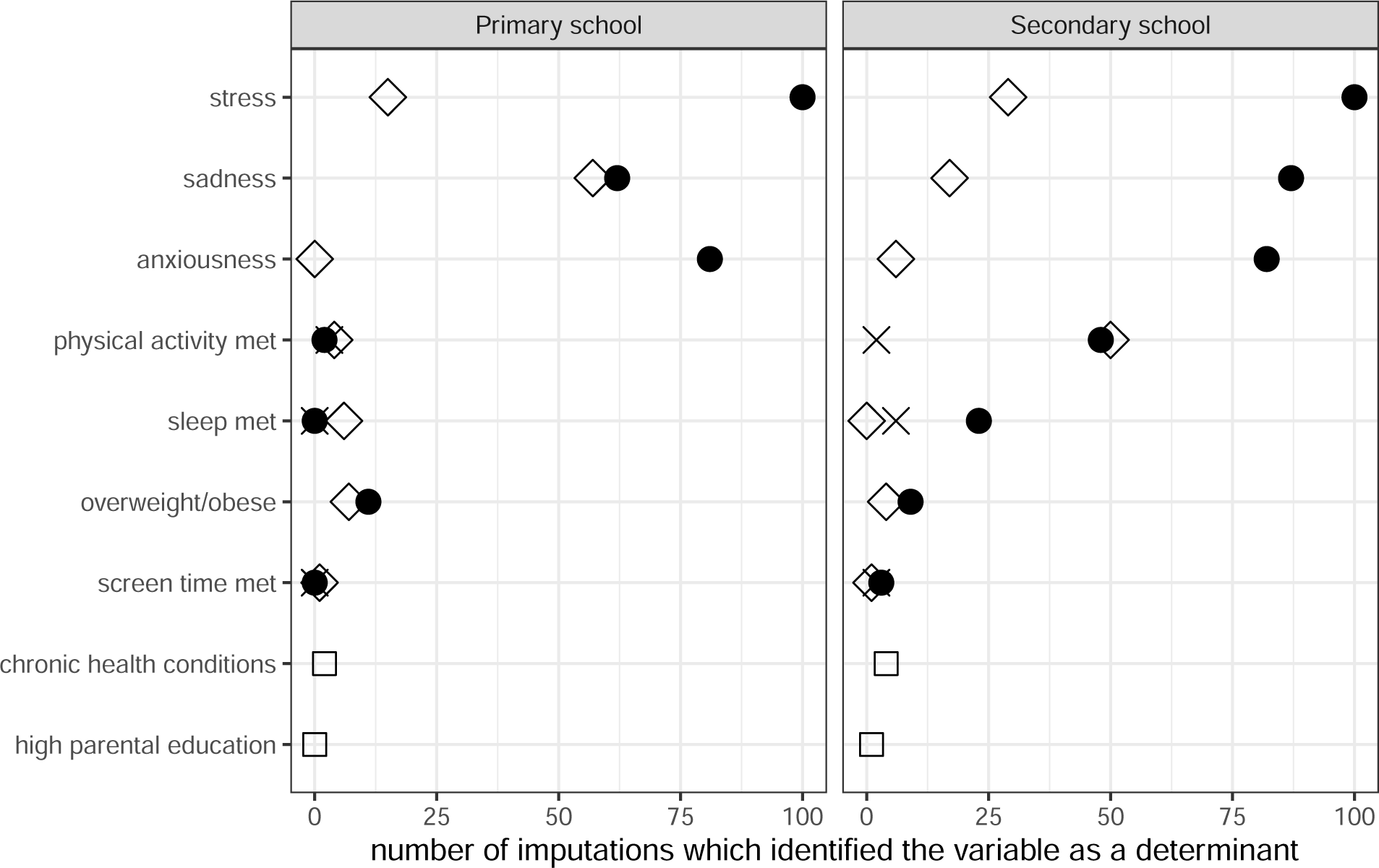
Variables identified as determinants of KINDL total scores by recursive partitioning after multiple imputation (with 100 imputations). More frequently identified variables are of greater importance than those identified in few imputations. For example, meeting screen time recommendations was identified in only a few imputations, and therefore did not appear to have a strong association with health-related quality of life, while meeting physical activity recommendations appeared to be of moderate importance related to health-related quality of life in secondary school students as it was identified in about 50/100 imputations.

For secondary school children, the models generally included stress, sadness, and anxiousness, with PA included in 50% of the models.

## Discussion

Psychological factors from 2021 such as stress, sadness, and anxiousness were most predictive of HRQOL in 2022, in both primary and secondary school children from a longitudinal cohort of schoolchildren during the COVID-19 pandemic from 2020-2022. Determinants from 2021 rather than 2022, explained a difference in KINDL total score of 13-15 points out of 100 in clusters of children and adolescents with or without these factors. Our study suggests that social and biological factors did not play a role in determining HRQOL, especially in primary school children. Only for adolescents in secondary school factors were such as PA, sleep, ST, chronic health conditions, and nationality identified as determinant of individual KINDL subscales. These results show that a major part of HRQOL in children and adolescents during the COVID-19 pandemic was explained by mental health determinants from periods when restrictions were still in place (in 2021).

The existence of sadness, anxiousness and stress as components of mental health led to a reduction in HRQOL of 12-15 points on a 0-100 scale in our sample, which is likely relevant from a public health perspective. To put this difference in context, it corresponds to or is even higher than the impact of family-based life changes, immigration status, or chronic health conditions such as asthma, headache, bipolar disorder or hemophilia [41–47]. Considering that Switzerland had experienced one of the mildest restrictions during the COVID-19 pandemic, and the high socio-economic status of our sample, the impact of the pandemic on mental health in children and adolescents and on their HRQOL is expected to be much larger in a more disadvantaged population and countries with more severe confinements [48].

A number of publications have examined the relationship between HRQOL and a small number of mostly single factors in children, primarily before the COVID-19 pandemic. It has been observed that parental education and family wealth [20], as well as cardiorespiratory fitness are correlated with HRQOL [15], along with PA [10–12], BMI [13], obesity [14] and ST [16]. Fewer studies have examined a broad range of potential factors and their association with HRQOL, and most were examined as single factors, especially since the onset of the pandemic [4,49,50]. These have identified psychological factors (self-esteem and emotions [19]), lifestyle factors (PA, sleep, ST, diet [18]), and sociodemographic factors (family education, poverty and race [51]; or unemployed parents, single parents, and non-western background [21]) as well as biological factors (disease burden, overweight, and chronic health conditions [21,51]). While sex in our analysis was not identified as an important determinant of HRQOL after considering other factors, other publications have observed significant univariate associations between HRQOL and sex [25,51,52]. Our analysis remains one of few studies that examined a broad range of possible determinants for HRQOL in a pandemic setting.

If sadness, anxiousness and stress are key determinants of HRQOL, the key public health implication is that reducing these three factors could improve HRQOL in children and adolescents. Improving mental health in children and adolescents likely requires individual, social and community strategies in order to be effective [53]. School-based interventions [54] could include self-help strategies [53,55], nature and green space [56–58] or strategies to improve lifestyle [16,59]. It may be that stress, anxiousness and sadness are only the nearest predictors in a pathway determining HRQOL. Changes to other factors may nevertheless improve mental health, thereby increasing HRQOL. It remains however unclear whether our findings can be generalized also to other settings beyond the pandemic.

This analysis has a number of strengths. It comprised a large sample (n = 1843) compared to similar studies on HRQOL, and was based on a sample from randomly selected schools for a whole canton that is representative or the general population of schoolchildren in the canton of Zurich and for health behaviors in Switzerland [60] (median participation rates within each class of 50% were high compared to similar studies [61], see also [62,63] as well as [64,65]). Prospective data was collected across 2 years during a pandemicduring which significant changes in life conditions throughout different levels of society (government, schools, families, peers) took place potentially affecting HRQOL. We based our concept on the bio-psycho-social health model, in line with a well-established conceptual model of HRQOL [7]. Stratification by age group accounted for differing behavior patterns and perceptions in primary school children versus adolescents in secondary school [52].

There are also several limitations. We did not measure HRQOL prior to the pandemic. While data collection for Ciao Corona did seek to examine changes in lifestyle and mental health during the COVID-19 pandemic, it did not *a priori* intend to explore determinants of HRQOL. Therefore, we have no information on some potentially interesting factors, for example mental status and/or substance abuse or lifestyle of parents. Depression and anxiety were not based on clinical criteria, but on single questions taken from the Health Behaviour in School-aged Children survey [66]. We cannot rule out some overlap between the KINDL items and the included potential determinants. Study participants were more likely to have Swiss nationality and more highly educated parents than the general population [67]. Had we been able to include also a more vulnerable, socially disadvantaged population, the study may have revealed an even stronger impact of mental health and other factors on HRQOL [20]. Additionally, data collection in our study leaned towards factors with possible negative impact [68,69]. Future studies may improve data collection by including a better balance between positive and negative factors.

In conclusion, we observed the psychological factors such as stress, sadness and anxiousness in 2021 were the main determinants of HRQOL in June 2022. Social and biological factors were generally not selected as determinants of overall HRQOL by our data-driven approach. A range of individual, family, community or school-based strategies are likely needed to improve mental health and consequently HRQOL in children and adolescents during such difficult pandemic-related times and beyond.

## Statements and Declarations

### Ethics approval and Informed Consent

The study was approved by the Ethics Committee of the Canton of Zurich, Switzerland (2020-01336). All participants provided written informed consent before being enrolled in the study.

### Data, Material and/or Code availability

The data used for this analysis can be obtained by request from the corresponding author.

### Authors’ contributions

SK and MAP initiated the project and preliminary design. SK, TR, SRH developed the design and methodology. SK, TR, AU, AR, SR, GPP, recruited study participants, collected, and managed the data. SRH performed statistical analysis and wrote the first draft of the manuscript. All authors contributed to the design of the study and interpretation of its results and revised and approved the manuscript for intellectual content. SK and SRH had access to and verified all underlying data. The corresponding author SK attests that all listed authors meet authorship criteria and that no others meeting the criteria have been omitted.

## Supporting information

Onine Resource 1

Online Resource 2

## Data Availability

The data used for this analysis can be obtained by request from the corresponding author.

## Acknowledgments

The authors thank Samuel Gunz for his assistance with preparing an earlier version of the HRQOL and lifestyle data used here.

## List of abbreviations

BMI: body mass index
HRQOL: health-related quality of life
IQR: interquartile range
PA: physical activity
ST: screen time

