## Supplementary material for "Determinants of health-related quality of life in healthy children and adolescents during the COVID-19 pandemic: results from a prospective longitudinal cohort study": Onine Resource 1

**Online Resource 1:** relevant questions from Ciao Corona questionnaires

Sarah R Haile, Gabriela P Peralta, Alessia Raineri, Sonja Rueegg, Agnė Ulytė, Milo A Puhan, Thomas Radtke, Susi Kriemler

### Contents

1 **Study characteristics** 1

2 **Demographic questions** 2

3 **Lifestyle** 3

4 **Mental Health** 4

5 **Symptoms compatible with post-Covid-19 condition** 4

6 **Characteristics of Parents and Family** 5

7 **Health-related quality of life** 7

**References** 8

### 1 Study characteristics

Ciao Corona was part of the Corona Immunitas research network, a program of 40 studies in Switzerland examinings seroprevalence of SARS-CoV-2 as well as related outcomes. Many questions included in the Ciao Corona surveys were the same as or very similar to those used in the other Corona Immunitas studies. The study design has been described previously [1,2].

Serological testing occurred at 5 timepoints:

- June/July 2020,
- October/November 2020,
- March 2021,
- November/December 2021, and
- June 2022.

Upon initial enrollment for serological testing, online **baseline questionnaires** were distributed to parents of the participating children by email to assess demographic characteristics, children's symptoms, possible SARS-CoV-2 infections and lifestyle behaviors. Parents were asked to fill the questionnaires together with their child.

Subsequently, parents and children were invited to complete further online **follow-up questionnaires** at the following timepoints:

- September/October 2020,
- January 2021,
- March 2021,
- September 2021,
- December 2021, and
- June 2022.

For each question, it is noted whether questions were included on "baseline" or "follow-up" form. If questions were introduced in later questionnaires, the specific timepoints are noted. The questions listed here are a subset of a much longer set of questions. Questions have been translated from German.

### 2 Demographic questions

**Date of birth** [baseline]

**Class year**

*What grade is the child in school?* [baseline]

- Possible responses: 1, 2, 3, 4, 5, 6, 7, 8, 9

**Sex of your child** [baseline]

- Possible responses: female, male, other/diverse

**Child's height**

*How tall is your son / daughter currently?* [baseline, follow-up September 2021, December 2021, June 2022]

**Child's weight**

*How much does your son / daughter weigh currently?* [baseline, follow-up September 2021, December 2021, June 2022]

Age-/sex-specific body mass index was compute from subjects' age, sex, height and weight according to [3]. Subjects were considered overweight if the body mass index was 1 or more standard deviations above the mean norm value.

**Chronic health conditions**

*Has a physician or medical specialist ever diagnosed your child with any of the following?*

- Possible responses (multiple answers possible): Asthma, Hay fever, Celiac, Lactose intolerance, allergies (other than hay fever), neurodermatitis/eczema, diabetes mellitus, Chronic intestinal inflammation (ulcerative colitis or Crohn's disease), high blood pressure, Attention deficit (ADHD, ADD), Epilepsy, Joint disease (e.g. arthritis), depression/anxiety, other disease [with comment field], I don't know, None

#### 3 Lifestyle

The wording of the following lifestyle questions was based on that found in previous studies of lifestyle in children and adolescents [4], and has been described previous for this study [5,6]. For sleep and screen time, we calculated a weighted average as follows: `[(weekday*5) + (weekend day*2)]/7`. The values were when used to determine adherence to international recommendations: $\geq 1$ hr/day of physical activity, $\leq 2$ hr/day of screen time [7], and age-specific sleep duration (10–13 hr/night for 5 years; 9–11 hr/night for 6–13 years; 8–10 hr/night for 14–16 years) [8].

**Physical activity**

*How many hours of sports or other physical activities per day (that caused sweating or increased breathing) did your child do on a typical weekday BEFORE the Corona epidemic (before March 16, 2020) (including school sports)?* [baseline June/July 2020]

*How many hours of sports or other physical activities per day (that caused sweating or increased breathing) did your child do on a typical weekend day BEFORE the Corona epidemic (before March 16, 2020) (including school sports)?* [baseline June/July 2020]

*How many hours of sports or other physical activities per day (that caused sweating or increased breathing) did your child do DURING the Corona epidemic (March 16 to May 11, 2020) (including school sports)?* [baseline June/July 2020]

*How many hours of sports or other physical activity per day (that caused sweating or increased breathing) does your son/daughter CURRENTLY do on a typical weekday (Monday to Friday)? Please include school sports.* [baseline November/December 2021, June 2022, follow-up]

*How many hours of sports or other physical activity per day (that caused sweating or increased breathing) does your son/daughter CURRENTLY do on a typical weekend day (Saturday, Sunday)?* [baseline November/December 2021, June 2022, follow-up]

**Screen time**

*How many hours per day did your child spend using electronic devices (e.g. cell phone, Playstation, Xbox, Nintendo, computer, TV) before the Corona epidemic (before March 16, 2020)? School lessons and schoolwork outside of school are not included.* [baseline June/July 2020]

*How many hours per day did your child spend using electronic devices (e.g. cell phone, Playstation, Xbox, Nintendo, computer, TV) during the Corona epidemic (period March 16, 2020 to May 11, 2020)? School lessons and schoolwork outside of school are not included.* [baseline June/July 2020]

*How many hours per day does your son/daughter CURRENTLY spend using electronic devices (e.g. cell phone, computer) for school on a typical weekday (Monday to Friday)?* [baseline November/December 2021, June 2022, follow-up September 2021, December 2021, June 2022]

*How many hours per day does your son/daughter CURRENTLY spend using electronic devices (e.g. cell phone, computer) for school on a typical weekend day (Saturday and Sunday)?* [baseline November/December 2021, June 2022, follow-up September 2021, December 2021, June 2022]

**Sleep**

*Before the Corona epidemic (before March 16, 2020), how many hours per day did your child sleep on a typical weekday?* [baseline June/July 2020]

*How many hours per day did your child sleep on a typical weekday during the Corona epidemic (March 16, 2020 to May 11, 2020)?* [baseline June/July 2020]

*How many hours per day does your son/daughter CURRENTLY sleep on a typical weekday (Monday to Friday)?* [baseline November/December 2021, June 2022, follow-up]

*How many hours per day does your son/daughter CURRENTLY sleep on a typical weekend day (Saturday and Sunday)?* [baseline November/December 2021, June 2022, follow-up]

### 4 Mental Health

*I felt sad, depressed* [baseline June/July 2020, November/December 2012, June 2022, follow-up]

- Possible responses: daily, several times a week, about once a week, about once a month, rarely or never

*I felt anxious, worried* [baseline June/July 2020, November/December 2012, June 2022, follow-up]

- Possible responses: daily, several times a week, about once a week, about once a month, rarely or never

*How would you rate the stress in your child's life right now?* [baseline November/December 2021, June 2022, follow-up]

- Possible responses: 1 (no stress), 2, 3, 4, 5, 6 (extreme stress)

### 5 Symptoms compatible with post-Covid-19 condition

*Has your child had any symptoms/complaints since fall break that have lasted at least 4 weeks? If NO, please select "No symptoms". If «YES», select the corresponding symptom(s).* [follow-up March 2021]

- possible responses [multiple possible]: no symptoms, fatigue, stuffy/runny nose, chest tightness, chest pain, cough, muscle pain, joint pain or swelling, headache, change sense of taste, changed sense of smell, difficulty concentrating, difficulty sleeping, increased need for sleep, weight loss, diarrhea, stomach pain, constipation, skin rash, palpitations, other [with comment field]

*Since the last survey, has your son/daughter had any symptoms/complaints that lasted at least 4 weeks (1 month)? If NO, please select "No symptoms". If YES, select the appropriate symptom(s).* [follow-up September 2021, December 2021]

- possible responses [multiple possible]: no symptoms, fatigue, stuffy/runny nose, chest tightness, chest pain, cough, muscle pain, joint pain or swelling, headache, change sense of taste, changed sense of smell, difficulty concentrating, difficulty sleeping, increased need for sleep, weight loss, diarrhea, stomach pain, constipation, skin rash, palpitations, other [with comment field]

*Since the last survey, has your son/daughter had any symptoms/complaints that lasted at least 4 weeks (1 month)? If NO, please select "No symptoms". If YES, select the appropriate symptom(s).* [follow-up June 2022]

- possible responses [multiple possible]: no symptoms, fatigue, stuffy/runny nose, chest tightness, chest pain, cough, wheezing, sore throat, muscle pain, joint pain or swelling, headache, dizziness, change sense of taste, changed sense of smell, difficulty concentrating, difficulty sleeping, mood swings, perception disorder, increased need for sleep, weight loss, diarrhea, stomach pain, abdominal pain, constipation, skin rash, palpitations, fever, other [with comment field]

For each of the checked above symptom responses, the following questions were additionally asked:

*How long has [this symptom] been present?*

- possible responses: 1-3 months, >3 months

*Is [this symptom] still present?*

- possible responses: yes / no

### 6 Characteristics of Parents and Family

**Parents' nationality**

*What is the nationality of the mother (the life partner of the father)?* [baseline]

- Possible responses (multiple answers possible): Switzerland, Germany, Austria, France, Italy, Spain, Portugal, Great Britain, USA, Turkey, Serbia, Montenegro, Kosovo, Croatia, other [with comment field]

*What is the nationality of the father (the life partner of the mother)?* [baseline]

- Possible responses (multiple answers possible): Switzerland, Germany, Austria, France, Italy, Spain, Portugal, Great Britain, USA, Turkey, Serbia, Montenegro, Kosovo, Croatia, other [with comment field]

**Parents' education**

*What is the mother's (father's partner)'s highest level of education?*

- Possible responses: No diploma, compulsory education, completed apprenticeship, college preparatory high school, vocational/technical university, university, don't know, other [with comment field] (Note: comments were also used to categorize all subjects who checked "other" into the above categories.)

*What is the father's (mother's partner)'s highest level of education?*

- Possible responses: No diploma, compulsory education, completed apprenticeship, college preparatory high school, vocational/technical university, university, don't know, other [with comment field] (Note: comments were also used to categorize all subjects who checked "other" into the above categories.)

**Change in employment situation**

*Have the working circumstances of the mother (the father's partner) currently changed compared to BEFORE the Corona crisis?* [baseline]

- Possible responses: yes, no

*If yes, specify:*

- Possible responses: You had temporarily stopped working or reduced your work, You had worked significantly more in the home office, You are currently on sick leave (with or without connection to corona infection), You are unemployed as a result of the corona crisis, Other

*Have the working circumstances of the father (the mother's partner) currently changed compared to BEFORE the Corona crisis?* [baseline]

- Possible responses: yes, no

*If yes, specify:*

- Possible responses: You had temporarily stopped working or reduced your work, You had worked significantly more in the home office, You are currently on sick leave (with or without connection to corona infection), You are unemployed as a result of the corona crisis, Other

*Father/mother's partner: Would you like to work more?* [follow-up September 2021, December 2021, June 2022]

- Possible responses: yes, no, not possible for health reasons, I would not like to answer this question

*Mother/father's partner Would you like to work more?* [follow-up September 2021, December 2021, June 2022]

- Possible responses: yes, no, not possible for health reasons, I would not like to answer this question

*Mother/father's partner: Looking back to 2019 (pre-pandemic period), has your employment changed due to the COVID-19 pandemic?* [follow-up September 2021, December 2021, June 2022]

- Possible responses: yes, no

*If yes, specify:* Increase in working hours, decrease in working hours, full or partial work from home, complete cessation of professional activity, medical leave, full or partial unemployment, termination of employment contract, non-renewal of the employment contract

*Father/mother's partner: Looking back to 2019 (pre-pandemic period), has your employment changed due to the COVID-19 pandemic?* [follow-up September 2021, December 2021, June 2022]

- Possible responses: yes, no

*If yes, specify:* Increase in working hours, decrease in working hours, full or partial work from home, complete cessation of professional activity, medical leave, full or partial unemployment, termination of employment contract, non-renewal of the employment contract

**Financial difficulties**

*Have your household experienced financial difficulties due to the change in employment?* [follow-up September 2021, December 2021, June 2022]

- possible responses: yes, no, I do not wish to answer this question.

### 7 Health-related quality of life

Health-related quality of life was assessed using the KINDL [9], Parent's Version for 7 to 17 year olds, at all baseline and follow-up timepoints. Computation of the KINDL total score is detailed in the manual.

Possible responses for all items are: never, rarely, sometimes, often, all the time.

**Physical Well-being**

During the past week …

1. … my child felt ill.
2. … my child had a headache or tummyache.
3. … my child was tired or worn out.
4. … my child felt strong or full of energy.

**Emotional Well-being**

During the past week …

1. … my child had fun and laughed a lot.
2. … my child didn't feel like doing anything.
3. … my child felt alone.
4. … my child felt scared or unsure of him/herself.

**Self-esteem**

During the past week …

1. … my child was proud of him/herself.
2. … my child felt on top of the world.
3. … my child felt pleased with him/herself.
4. … my child had lots of good ideas.

**Family**

During the past week …

1. … my child got on well with us as parents.
2. … my child felt fine at home.

3. … we quarrelled at home.
4. … my chil felt that I was bossing him/her around.

**Friends**

During the past week …

1. … my child did things together with friends.
2. … my child was liked by other kids.
3. … my child got along well with his/her friends.
4. … my child felt different from other children.

**School**

During the past week …

1. … my child easily coped with schoolwork.
2. … my child enjoyed the school lessons.
3. … my child worried about his/her future.
4. … my child was afraid about bad marks or grades.
