## Supplementary material for "Determinants of health-related quality of life in healthy children and adolescents during the COVID-19 pandemic: results from a prospective longitudinal cohort study": Online Resource 2

**Online Resource 2:** Further details on pandemic restrictions in Switzerland, detailed statistical methods, and additional tables and figures.

Sarah R Haile, Gabriela P Peralta, Alessia Raineri, Sonja Rueegg, Agnė Ulytė, Milo A Puhan, Thomas Radtke, Susi Kriemler

### Contents

**1 Pandemic restrictions in Switzerland** **1**

**2 Detailed Statistical Methods** **1**

**References** **2**

**3 Supplementary Figures and Tables** **4**
3.1 Details for Figures S4-S9 . . . . . . . . . . . . . . . . . . . . . . . . . . . . . . . . 4
3.2 Details for Figures S10-S15 . . . . . . . . . . . . . . . . . . . . . . . . . . . . . . 4

**Computational Details** **15**

### 1 Pandemic restrictions in Switzerland

Pandemic restrictions in Switzerland included a brief lockdown from March to May 2020 during which schools were completely remote [1], with a partial return to in-person school (e.g. with smaller classes or hybrid in person and online) until July 2020 and full in-person attendance thereafter. In addition, many activities, such as school trips, musical events, and sporting events or training, were either entirely cancelled or were conducted online. Despite quite high case rates [2], schoolchildren in Switzerland remained at school with additional regional regulations which included mask wearing (primary school from Jan. 21, 2021 for 4th graders and above or Dec. 9, 2021 for 1st graders and older to Feb. 20, 2022, secondary school from Oct. 28, 2020 to Feb. 20, 2022), quarantine and isolation measures (10 day requirement until Jan. 12, 2022, then 5 day requirement until March 30, 2022) and weekly pooled testing in some, but not all schools (from August 2021 to Feb. 20, 2022). Other activities, such as extracurricular sports, continued with the use of masks, small groups, availability only to children under the age of 16, and other restrictions through March 2021. Thereafter, most activities both in and outside of school continued without restrictions for children and adolescents. The situation in Switzerland during the pandemic provides an opportunity to assess changes in lifestyle and HRQOL where preventive measures for children were relatively mild compared to other countries [3].

### 2 Detailed Statistical Methods

We used conditional inference trees [4,5] estimated by binary recursive partitioning to identify possible determinants of health-related quality of life (HRQOL) in children and adolescents. These models seek to make homogeneous subgroups, i.e. clusters, of the sample with respect to the outcome of interest. Generally, the algorithm 1) searches for the variable with the strongest association to the outcome, and then 2) splits the values of that variable into two groups, and repeats this process until some stopping criteria (in this analysis, p-value < 0.05 or sample size < 25) are reached. While all types of variables can be selected in step 1 of this algorithm, categorical and continuous variables have more flexibility in terms of selected thresholds in step 2 than binary variables do. Conditional inference trees select variables in an unbiased manner, without being affected by overfitting [4]. Such models identify subgroups defined by combinations of covariates, without needing to *a priori* specify interaction terms or consider multicollinearity [6], and thresholds do not need to be prespecified. Standard regression models, even when combined with model selection, do not identify homogenous subgroups. This procedure was repeated for the KINDL total score as well as for each of its subscales, and stratified by age group.

Due the presence of missing observations in some of the variables, we employed so-called surrogate splits to account for this missingness without excluding subjects [4]. As a sensitivity analysis, multiple imputation using chained equations was used to impute missing covariates [7] (m = 100), and then recursive partitioning was used to identify significant predictors of HRQOL in each of the imputed datasets. We then counted how often each variable was included in the model selection procedure, deeming those that appeared in more than 50% of the models to be important determinants.

The conditional inference trees were fit using `ctree` from the R package `partykit` [4,8]. Missing covariate information was handled by `ctree` directly (or in the sensitvity analysis, with multiple imputation). Multiple imputation was performed using the `mice` package [9]. All analysis was performed in R (R version 4.3.2 (2023-10-31)).

### 3 Supplementary Figures and Tables

S7 Model results for KINDL family scale in primary school children. See Section 3.1. 12
S8 Model results for KINDL friends scale in primary school children. See Section 3.1. 13
S9 Model results for KINDL school scale in primary school children. See Section 3.1. 14
S10 Model results for KINDL physical scale in secondary school children. See Section 3.2. . . . . . . . . . . . . . . . . . . . . . . . . . . . . . . . . . . . . . . . 16
S11 Model results for KINDL emotional scale in secondary school children. See Section 3.2. . . . . . . . . . . . . . . . . . . . . . . . . . . . . . . . . . . . . . 17
S12 Model results for KINDL selfesteem scale in secondary school children. See Section 3.2. . . . . . . . . . . . . . . . . . . . . . . . . . . . . . . . . . . . . . 18
S13 Model results for KINDL family scale in secondary school children. See Section 3.2. . . . . . . . . . . . . . . . . . . . . . . . . . . . . . . . . . . . . . . . 19
S14 Model results for KINDL friends scale in secondary school children. See Section 3.2. . . . . . . . . . . . . . . . . . . . . . . . . . . . . . . . . . . . . . . . 20
S15 Model results for KINDL school scale in secondary school children. See Section 3.2. 21

##### 3.1 Details for Figures S4-S9

Recursive partitioning tree for HRQOL (KINDL total score) in 2022 in primary school children. Identified determinants are stress, sadness and anxiousness in 2021, as well as chronic health conditions (`chronic`), Swiss nationality, and self-rated health. Sadness and anxiousness could have occurred once per month (1M), once per week (1W), more than once per week (>1W) or daily (D). Stress was considered on a 5-point scale from 1 (no stress) to 4 (high stress). Self-rated health was rated as excellent, good, moderately good or bad. Other variables included in the model could not be used to create more homogeneous groups with respect to KINDL total score. For each subgroup, median KINDL total score, interquartile range (IQR), mean ± standard deviation and sample size (n) are given.

##### 3.2 Details for Figures S10-S15

Recursive partitioning tree for health-related quality of life (HRQOL, KINDL total score and its subscales) in 2022 in secondary school children. Identified determinants are stress, sadness and anxiousness in 2021, as well as sex, chronic health conditions (`chronic`), Swiss nationality, whether physical activity recommendations were met (`PA met`), whether sleep recommendations were met (`sleep met`) and self-rated health. Sadness and anxiousness could have occurred once per month (1M), once per week (1W), more than once per week (>1W) or daily (D). Stress was considered on a 5-point scale from 1 (no stress) to 4 (high stress). Self-rated health was rated as excellent, good, moderately good or bad. Other variables included in the model could not be used to create more homogeneous groups with respect to KINDL total score. For each subgroup, median KINDL total score, interquartile range (IQR), mean ± standard deviation and sample size (n) are given.

Table S1: Median and interquartile range for KINDL total score by year and age group.

| grp | 2020 | 2021 | 2022 |
|---|---|---|---|
| primary | 82.3 [77.1 - 86.5] | 81.2 [75.0 - 86.5] | 80.2 [74.0 - 85.4] |
| secondary | 79.2 [72.9 - 85.1] | 76.0 [68.8 - 82.6] | 74.0 [67.7 - 81.2] |

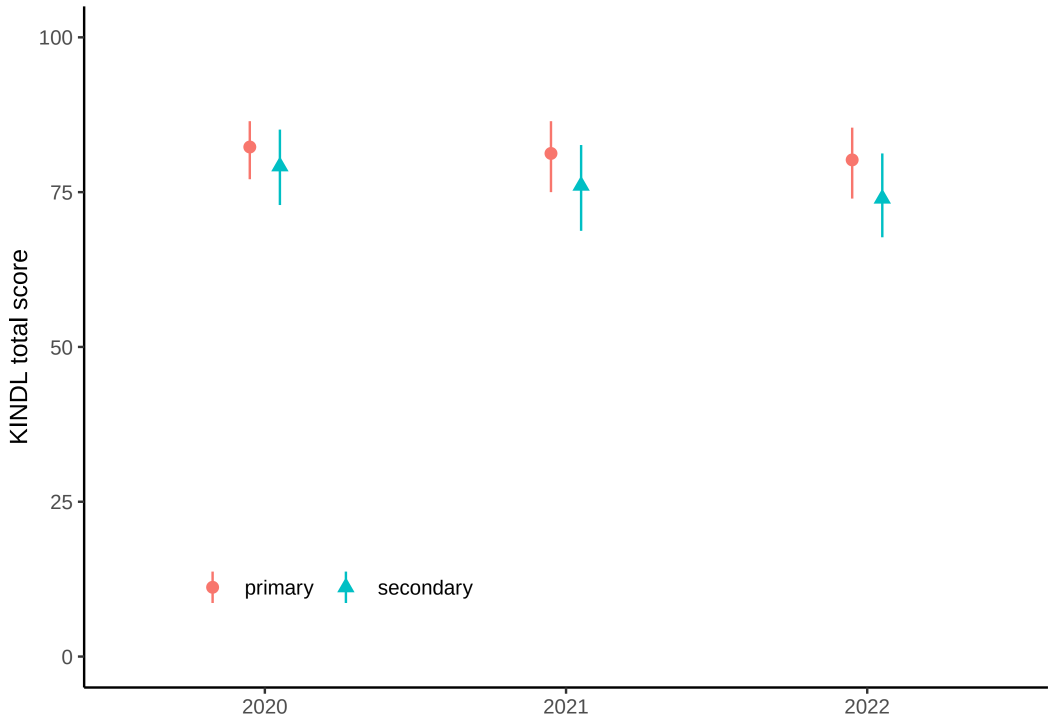

Figure S1: Median and interquartile range for KINDL total score by year and age group.

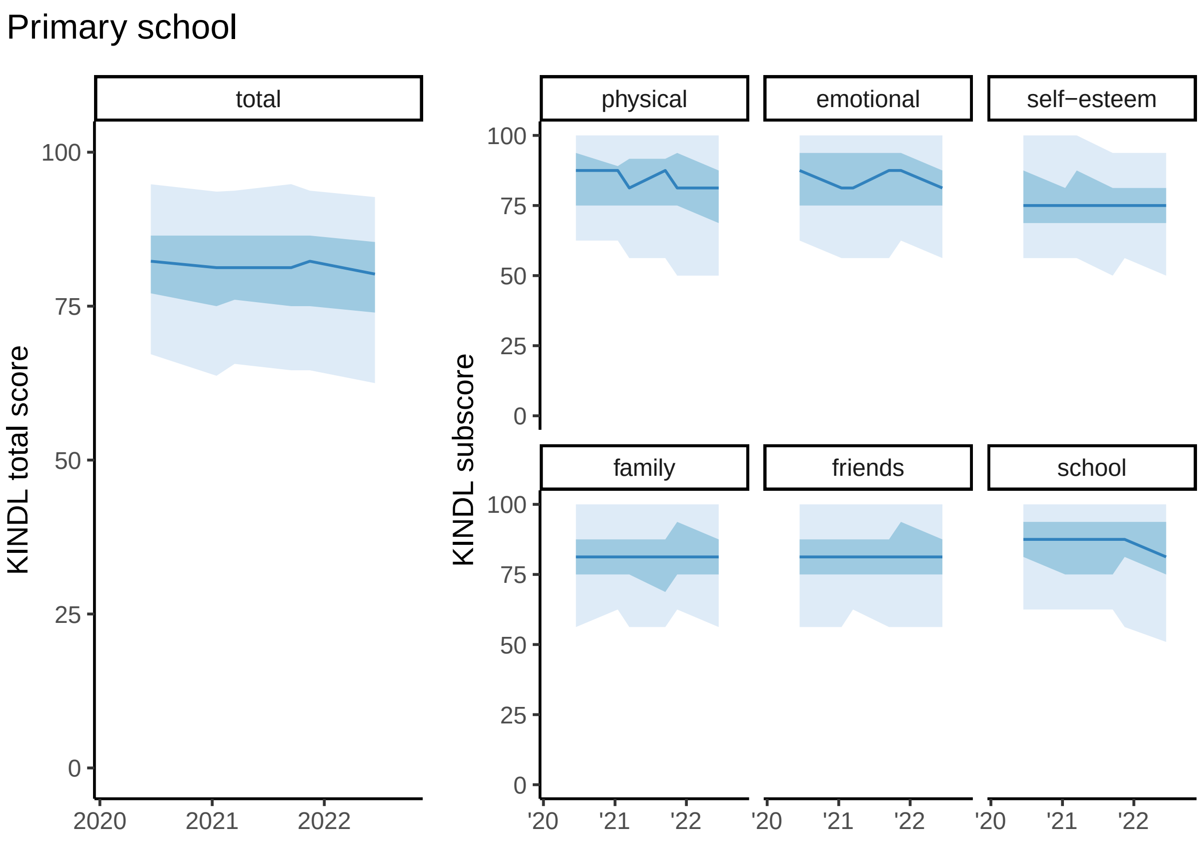

Figure S2: Changes over time in KINDL total score and its subscales, in primary school. Median (line), interquartile range (dark blue region) and 5th to 95th percentile (light blue region) are shown.

Table S2: Median and IQR for KINDL total score and its subscales by timepoint, for primary school participants.

| subscale | 06/2020 | 01/2021 | 03/2021 | 09/2021 | 11/2021 | 06/2022 |
|---|---|---|---|---|---|---|
| total | 82.3 (77.1-86.5) | 81.2 (75.0-86.5) | 81.2 (76.0-86.5) | 81.2 (75.0-86.5) | 82.3 (75.0-86.5) | 80.2 (74.0-85.4) |
| physical | 87.5 (75.0-93.8) | 87.5 (75.0-89.1) | 81.2 (75.0-91.7) | 87.5 (75.0-91.7) | 81.2 (75.0-93.8) | 81.2 (68.8-87.5) |
| emotional | 87.5 (75.0-93.8) | 81.2 (75.0-93.8) | 81.2 (75.0-93.8) | 87.5 (75.0-93.8) | 87.5 (75.0-93.8) | 81.2 (75.0-87.5) |
| self-esteem | 75.0 (68.8-87.5) | 75.0 (68.8-81.2) | 75.0 (68.8-87.5) | 75.0 (68.8-81.2) | 75.0 (68.8-81.2) | 75.0 (68.8-81.2) |
| family | 81.2 (75.0-87.5) | 81.2 (75.0-87.5) | 81.2 (75.0-87.5) | 81.2 (68.8-87.5) | 81.2 (75.0-93.8) | 81.2 (75.0-87.5) |
| friends | 81.2 (75.0-87.5) | 81.2 (75.0-87.5) | 81.2 (75.0-87.5) | 81.2 (75.0-87.5) | 81.2 (75.0-93.8) | 81.2 (75.0-87.5) |
| school | 87.5 (81.2-93.8) | 87.5 (75.0-93.8) | 87.5 (75.0-93.8) | 87.5 (75.0-93.8) | 87.5 (81.2-93.8) | 81.2 (75.0-93.8) |

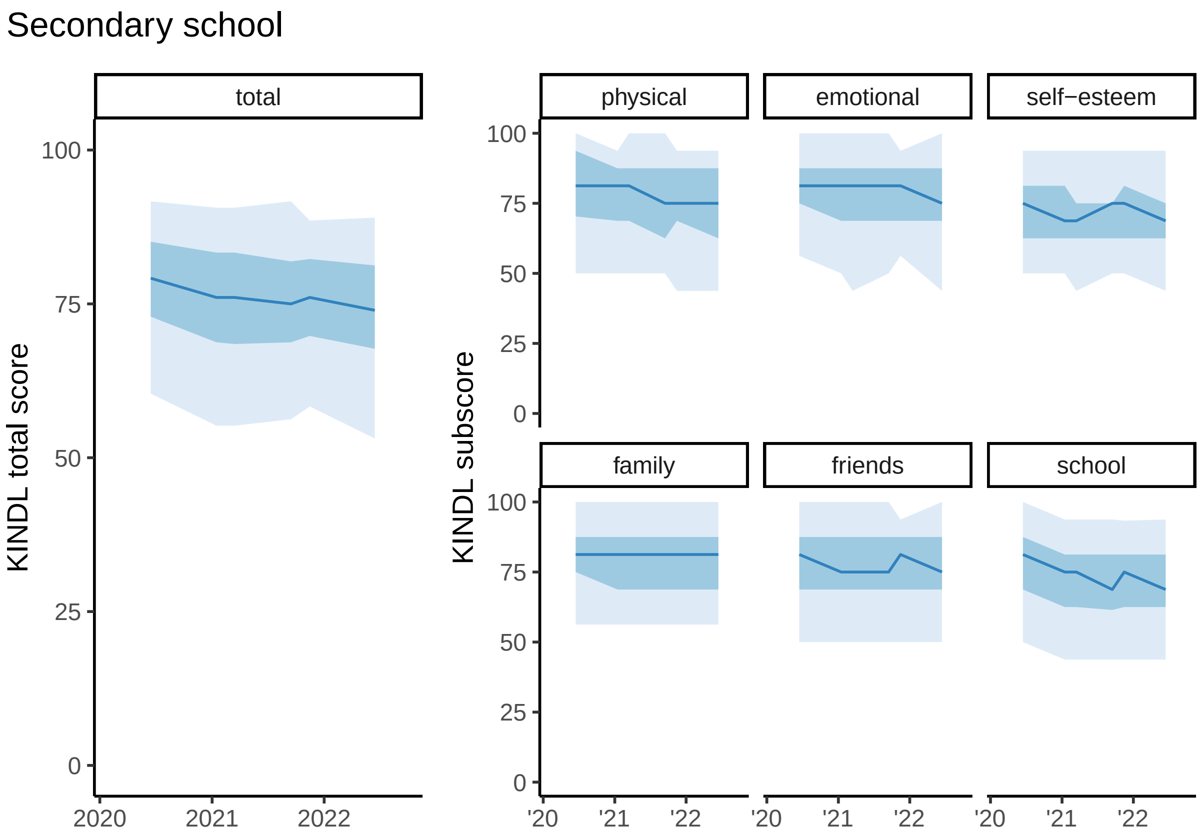

Figure S3: Changes over time in KINDL total score and its subscales, in secondary school. Median (line), interquartile range (dark blue region) and 5th to 95th percentile (light blue region) are shown.

Table S3: Median and IQR for KINDL total score and its subscales by timepoint, for secondary school participants.

| subscale | 06/2020 | 01/2021 | 03/2021 | 09/2021 | 11/2021 | 06/2022 |
|---|---|---|---|---|---|---|
| total | 79.2 (72.9-85.1) | 76.0 (68.8-83.3) | 76.0 (68.5-83.3) | 75.0 (68.8-81.9) | 76.0 (69.8-82.3) | 74.0 (67.7-81.2) |
| physical | 81.2 (70.3-93.8) | 81.2 (68.8-87.5) | 81.2 (68.8-87.5) | 75.0 (62.5-87.5) | 75.0 (68.8-87.5) | 75.0 (62.5-87.5) |
| emotional | 81.2 (75.0-87.5) | 81.2 (68.8-87.5) | 81.2 (68.8-87.5) | 81.2 (68.8-87.5) | 81.2 (68.8-87.5) | 75.0 (68.8-87.5) |
| self-esteem | 75.0 (62.5-81.2) | 68.8 (62.5-81.2) | 68.8 (62.5-75.0) | 75.0 (62.5-75.0) | 75.0 (62.5-81.2) | 68.8 (62.5-75.0) |
| family | 81.2 (75.0-87.5) | 81.2 (68.8-87.5) | 81.2 (68.8-87.5) | 81.2 (68.8-87.5) | 81.2 (68.8-87.5) | 81.2 (68.8-87.5) |
| friends | 81.2 (68.8-87.5) | 75.0 (68.8-87.5) | 75.0 (68.8-87.5) | 75.0 (68.8-87.5) | 81.2 (68.8-87.5) | 75.0 (68.8-87.5) |
| school | 81.2 (68.8-87.5) | 75.0 (62.5-81.2) | 75.0 (62.5-81.2) | 68.8 (61.5-81.2) | 75.0 (62.5-81.2) | 68.8 (62.5-81.2) |

Table S4: Results for KINDL total score from recursive partitioning after multiple imputation in primary school participants. Number $n$ indicates how many imputations, out of 100, that a variable was identified by the recursive partitioning model. BMI indicates body mass index.

| term | demographic | 2019 | 2020 | 2021 |
|---|---|---|---|---|
| sadness | | 0 | 57 | 62 |
| stress | | 0 | 15 | 100 |
| anxiousness | | 0 | 0 | 81 |
| overweight/obese | | 0 | 7 | 11 |
| physical activity met | | 3 | 4 | 2 |
| sleep met | | 0 | 6 | 0 |
| chronic conditions | 2 | 0 | 0 | 0 |
| screen time met | | 0 | 1 | 0 |
| parents education | 0 | 0 | 0 | 0 |

Table S5: Results for KINDL total score from recursive partitioning after multiple imputation in secondary school participants. Number $n$ indicates how many imputations, out of 100, that a variable was identified by the recursive partitioning model. BMI indicates body mass index.

| term | demographic | 2019 | 2020 | 2021 |
|---|---|---|---|---|
| stress | | 0 | 29 | 100 |
| sadness | | 0 | 17 | 87 |
| physical activity met | | 2 | 50 | 48 |
| anxiousness | | 0 | 6 | 82 |
| sleep met | | 6 | 0 | 23 |
| overweight/obese | | 0 | 4 | 9 |
| screen time met | | 2 | 1 | 3 |
| chronic conditions | 4 | 0 | 0 | 0 |
| parents education | 1 | 0 | 0 | 0 |

Table S6: Summary of model results for KINDL total score and its subscales in primary school children (included 1, not included 0).

| vars | KINDL | physical | emotional | self-esteem | family | friends | school |
|---|---|---|---|---|---|---|---|
| sadness 2021 | 1 | 1 | 1 | 0 | 0 | 0 | 1 |
| anxiousness 2021 | 1 | 0 | 0 | 1 | 0 | 1 | 0 |
| stress 2021 | 1 | 1 | 1 | 1 | 1 | 1 | 1 |
| sex | 0 | 0 | 0 | 1 | 0 | 0 | 0 |
| chronic | 0 | 0 | 0 | 0 | 0 | 1 | 0 |

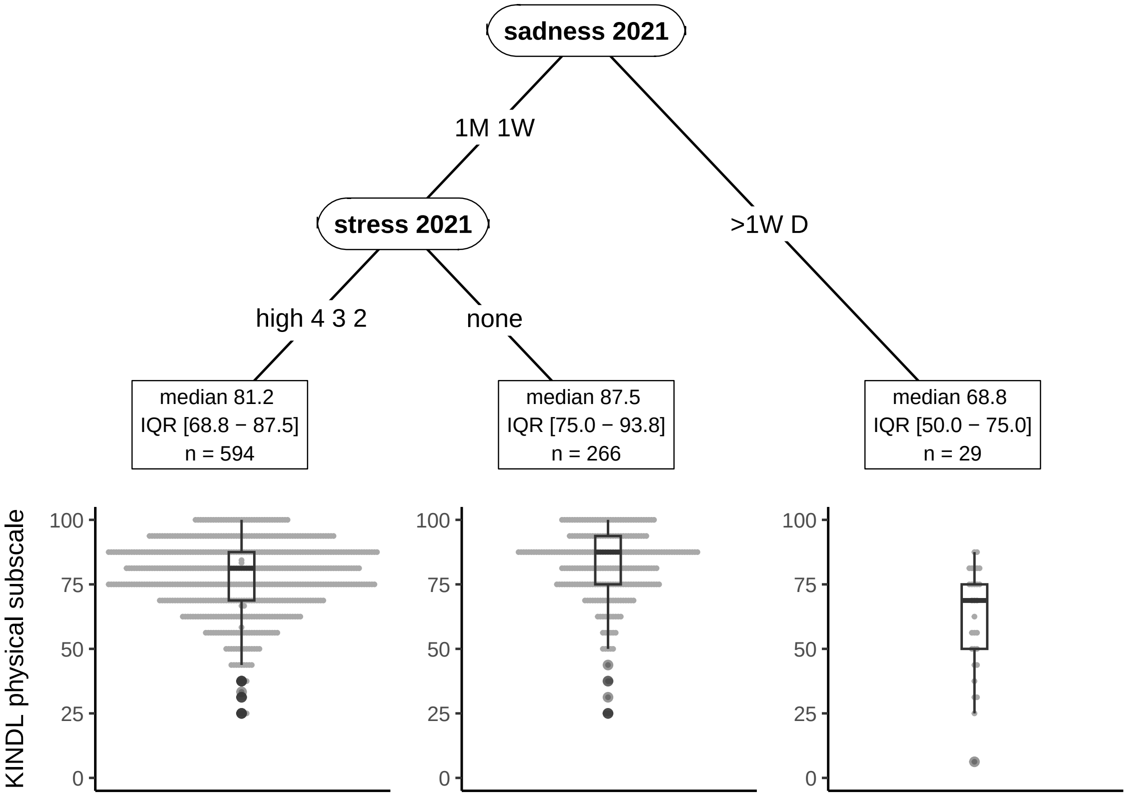

Figure S4: Model results for KINDL physical scale in primary school children. See Section 3.1.

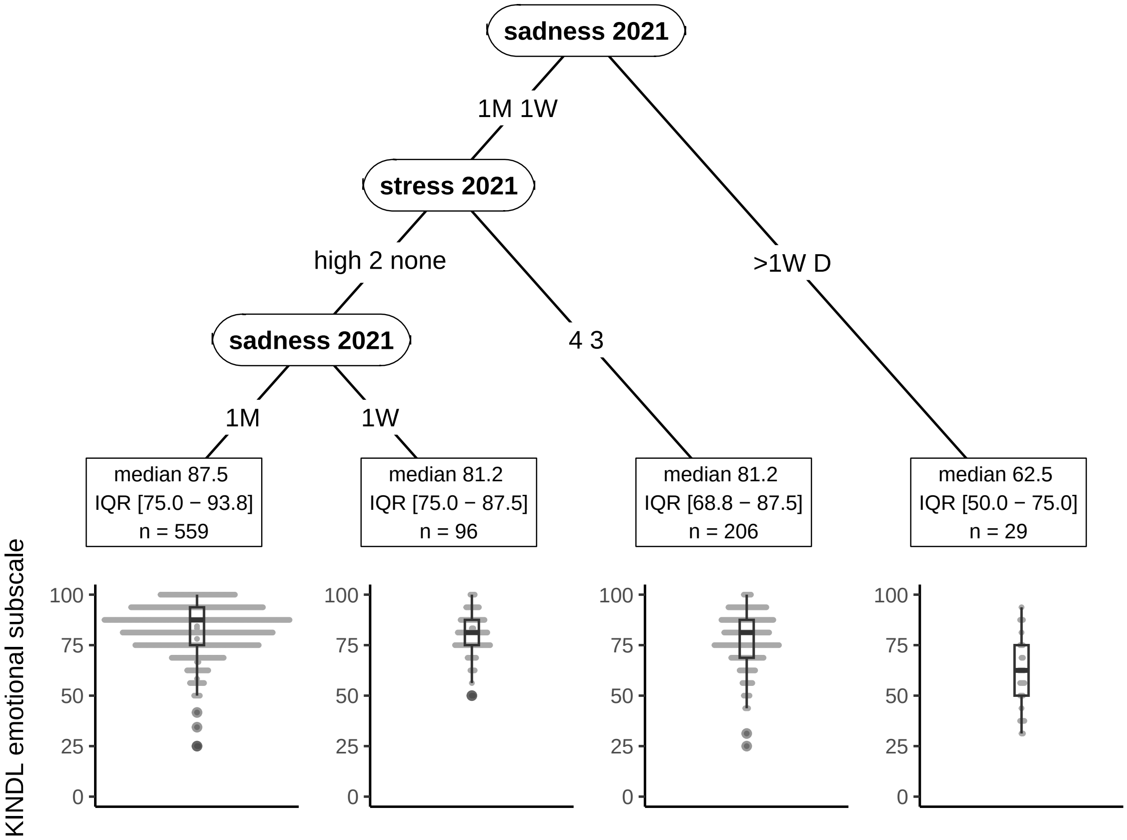

Figure S5: Model results for KINDL emotional scale in primary school children. See Section 3.1.

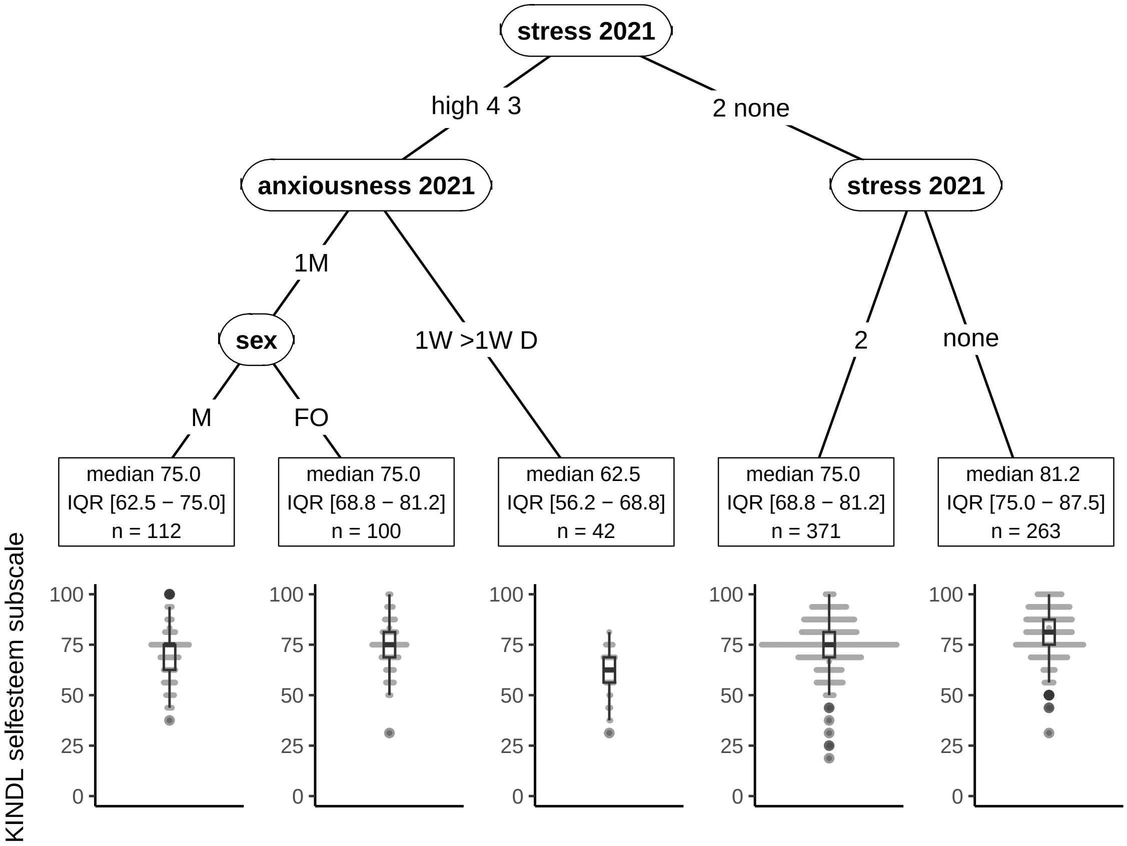

Figure S6: Model results for KINDL selfesteem scale in primary school children. See Section 3.1.

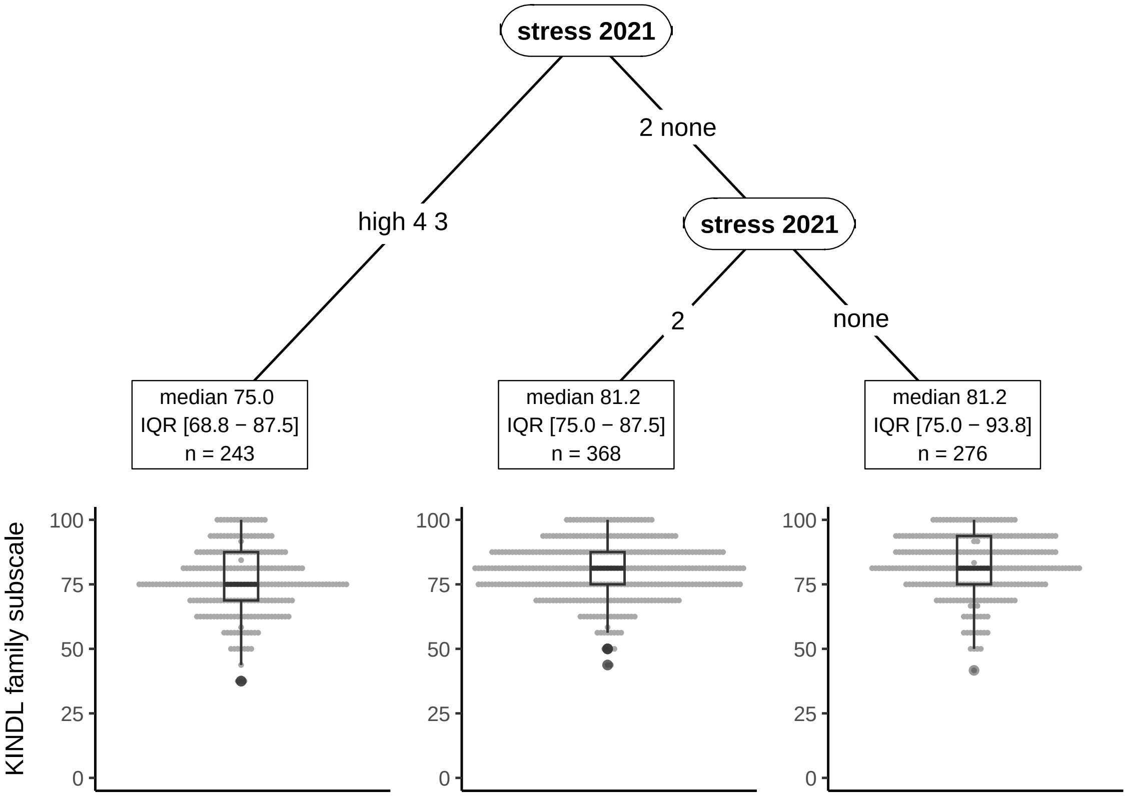

Figure S7: Model results for KINDL family scale in primary school children. See Section 3.1.

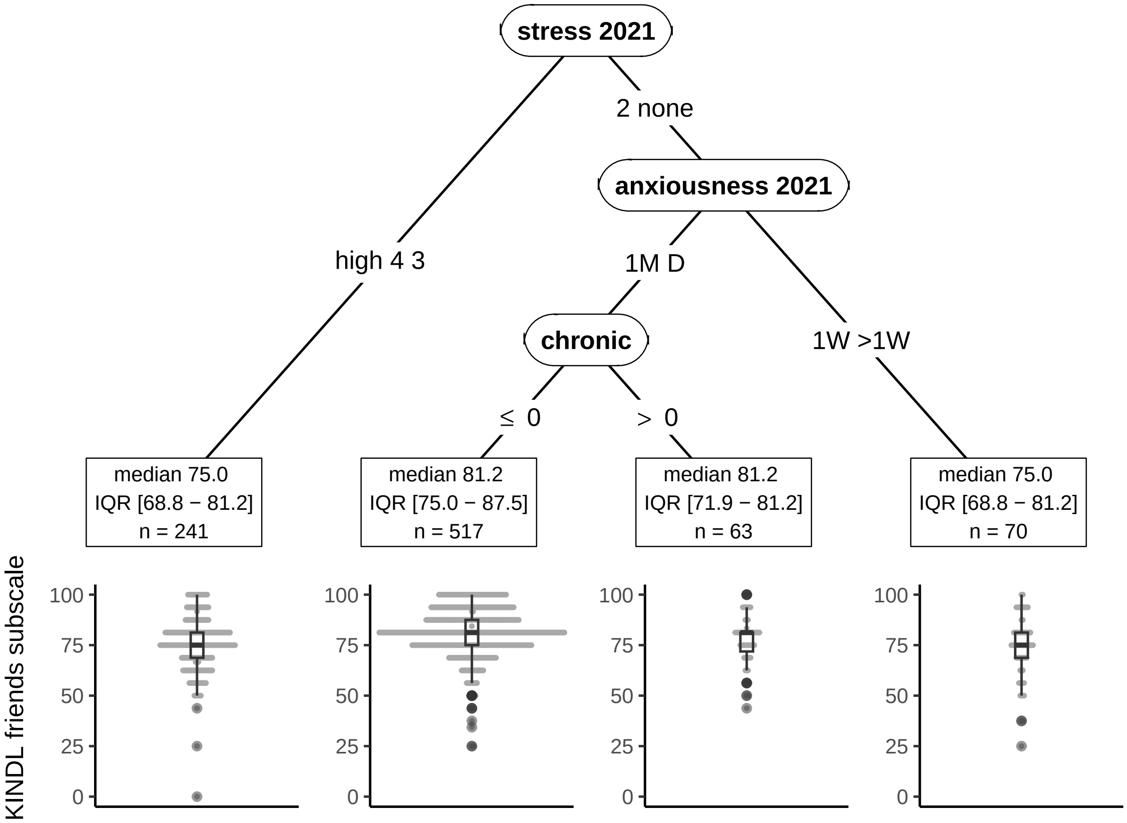

Figure S8: Model results for KINDL friends scale in primary school children. See Section 3.1.

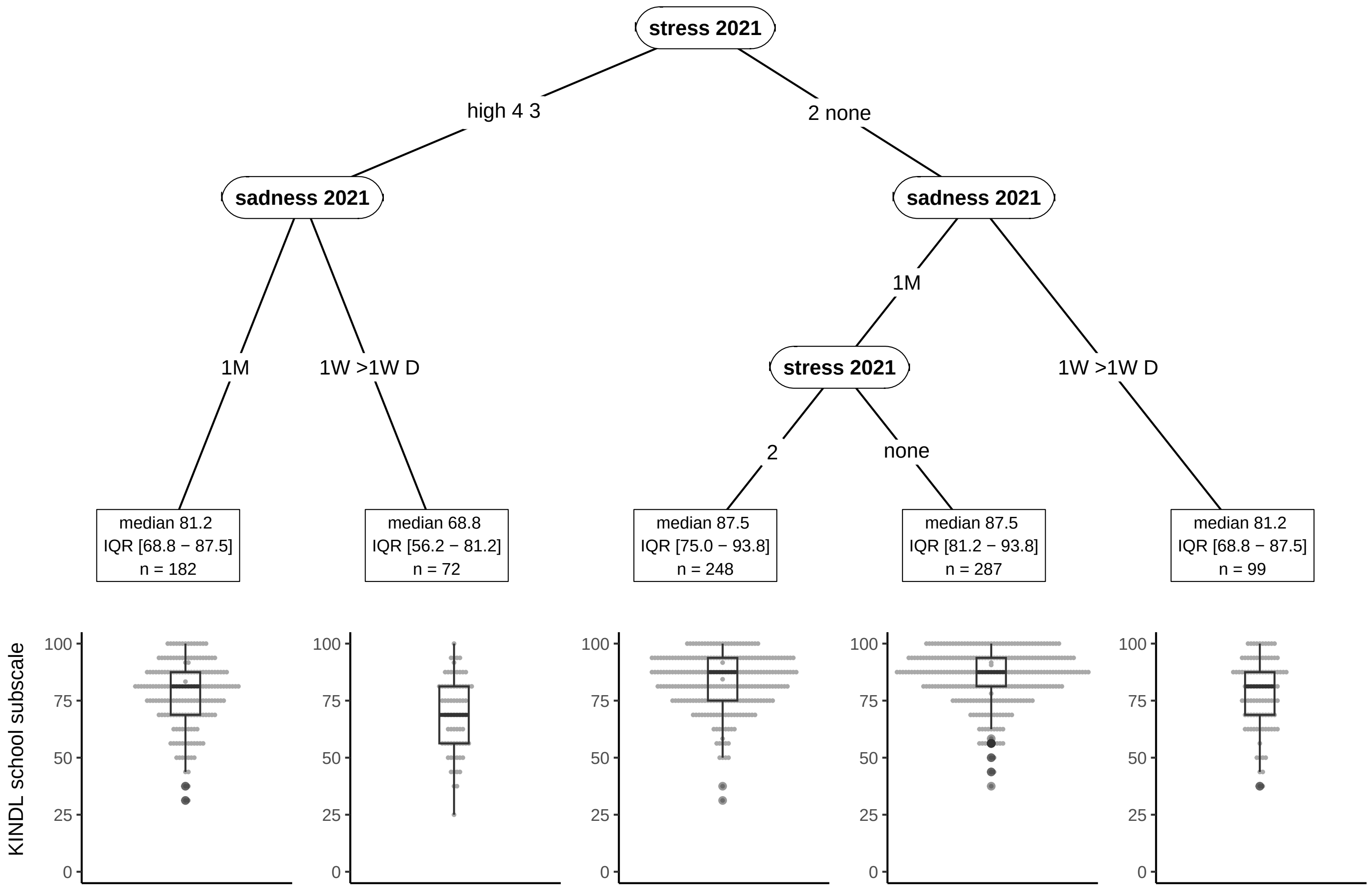

Figure S9: Model results for KINDL school scale in primary school children. See Section 3.1.

Table S7: Summary of model results for KINDL total score and its subscales in secondary school children (included 1, not included 0).

| vars | KINDL | physical | emotional | self-esteem | family | friends | school |
|---|---|---|---|---|---|---|---|
| sadness 2021 | 1 | 1 | 1 | 1 | 0 | 1 | 0 |
| anxiousness 2021 | 1 | 0 | 1 | 0 | 0 | 1 | 1 |
| stress 2021 | 1 | 1 | 1 | 1 | 1 | 0 | 1 |
| sex | 0 | 1 | 0 | 0 | 0 | 0 | 0 |
| sleep met 2020 | 0 | 0 | 1 | 0 | 0 | 0 | 0 |
| PA met 2021 | 0 | 0 | 0 | 1 | 0 | 1 | 0 |
| sleep met 2021 | 0 | 0 | 0 | 0 | 1 | 0 | 0 |
| ST met 2020 | 0 | 0 | 0 | 0 | 0 | 1 | 0 |
| chronic | 0 | 0 | 0 | 0 | 0 | 1 | 0 |
| stress 2020 | 0 | 0 | 0 | 0 | 0 | 0 | 1 |
| ST met 2021 | 0 | 0 | 0 | 0 | 0 | 0 | 1 |

### Computational Details

- R version: R version 4.3.2 (2023-10-31)
- Base packages: grid, stats, graphics, grDevices, utils, datasets, methods, base
- Other packages: abe 3.0.1, patchwork 1.1.3, mice 3.16.0, knitr 1.45, corrplot 0.92, rlang 1.1.2, ggparty 1.0.0, partykit 1.2.20, mvtnorm 1.2.3, libcoin 1.0.9, gtsummary 1.7.2, lubridate 1.9.2, forcats 1.0.0, stringr 1.5.1, dplyr 1.1.3, purrr 1.0.2, readr 2.1.4, tidyr 1.3.0, tibble 3.2.1, ggplot2 3.4.3, tidyverse 2.0.0

This document was generated on 2024-01-16 at 17:16.

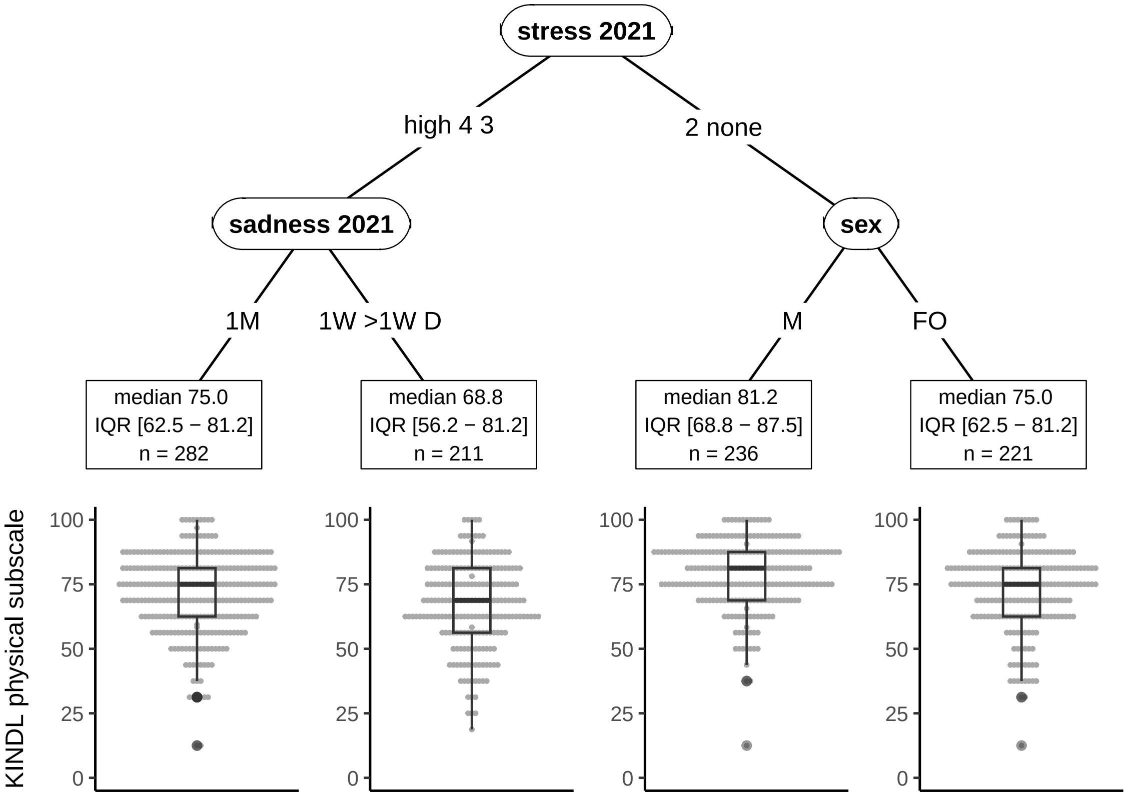

Figure S10: Model results for KINDL physical scale in secondary school children. See Section 3.2.

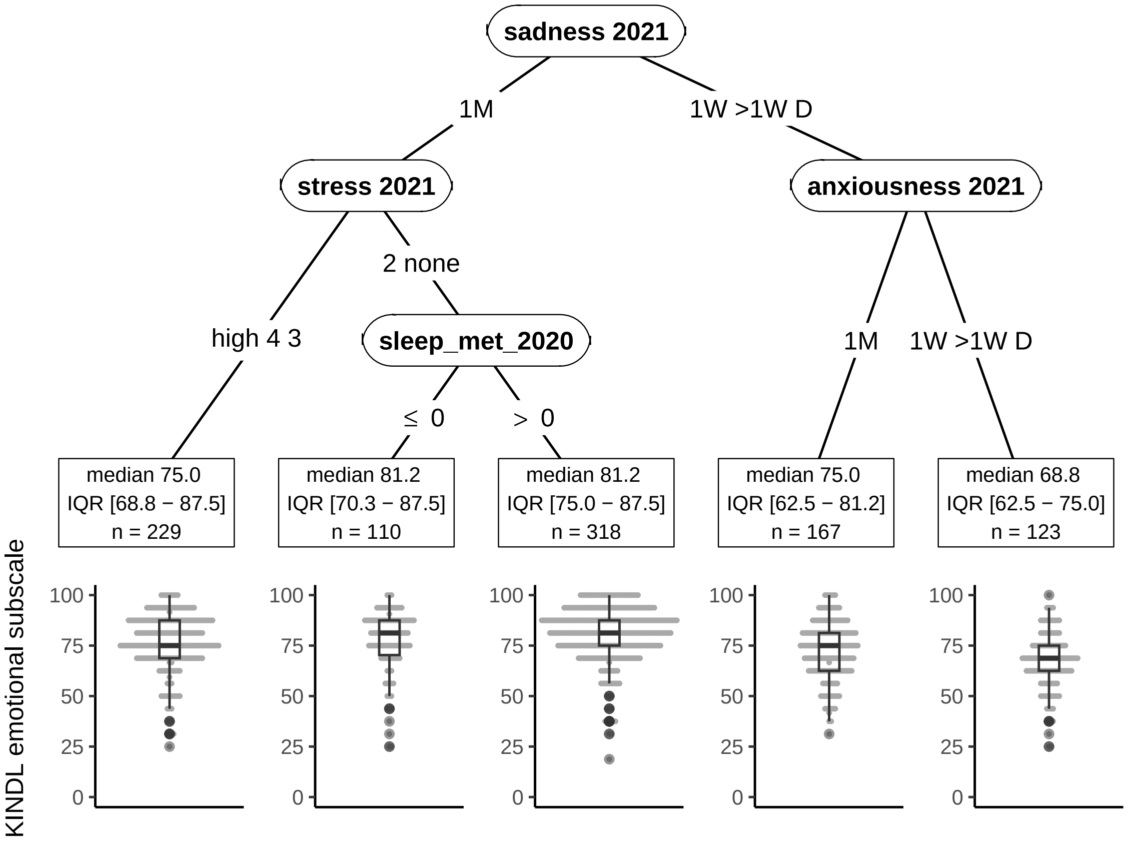

Figure S11: Model results for KINDL emotional scale in secondary school children. See Section 3.2.

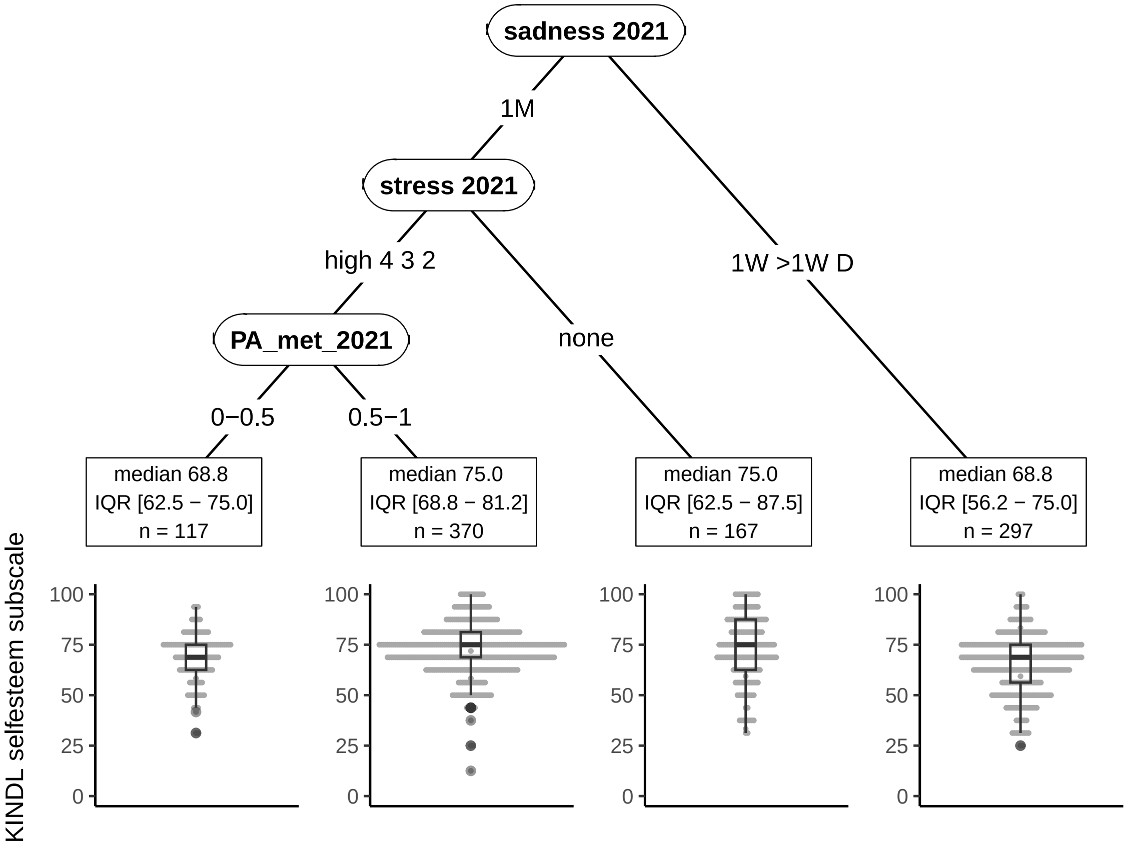

Figure S12: Model results for KINDL selfesteem scale in secondary school children. See Section 3.2.

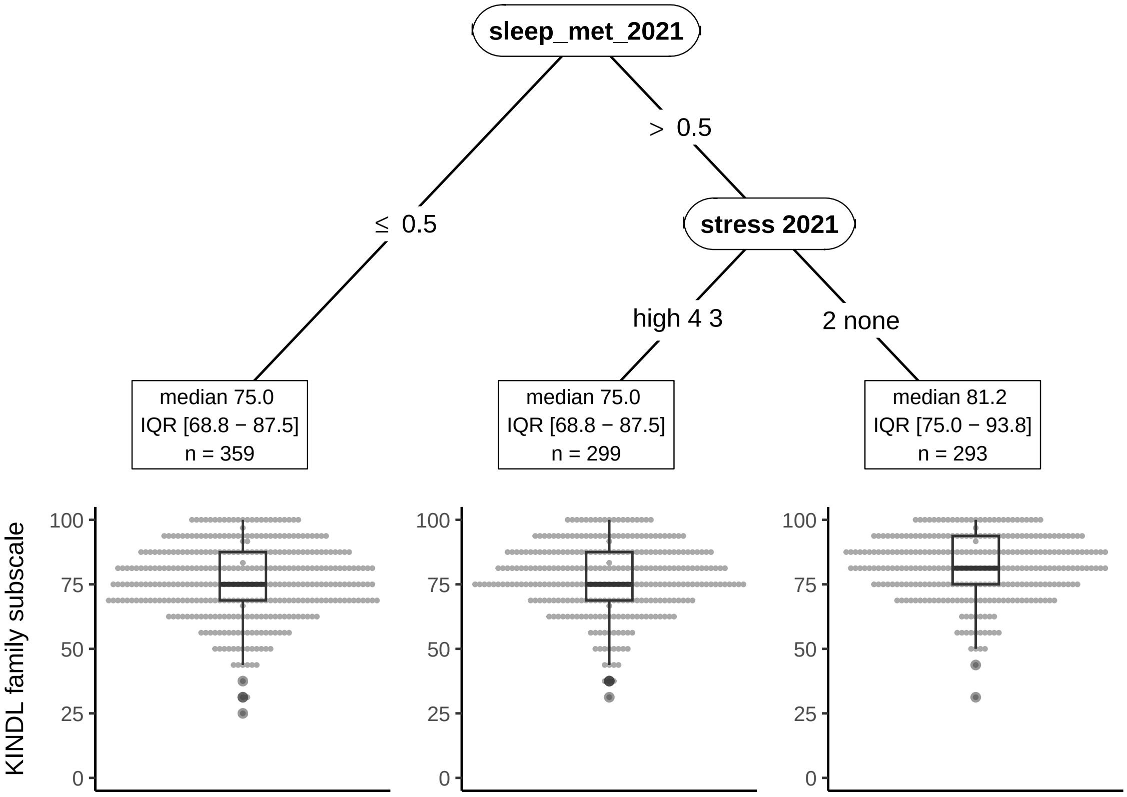

Figure S13: Model results for KINDL family scale in secondary school children. See Section 3.2.

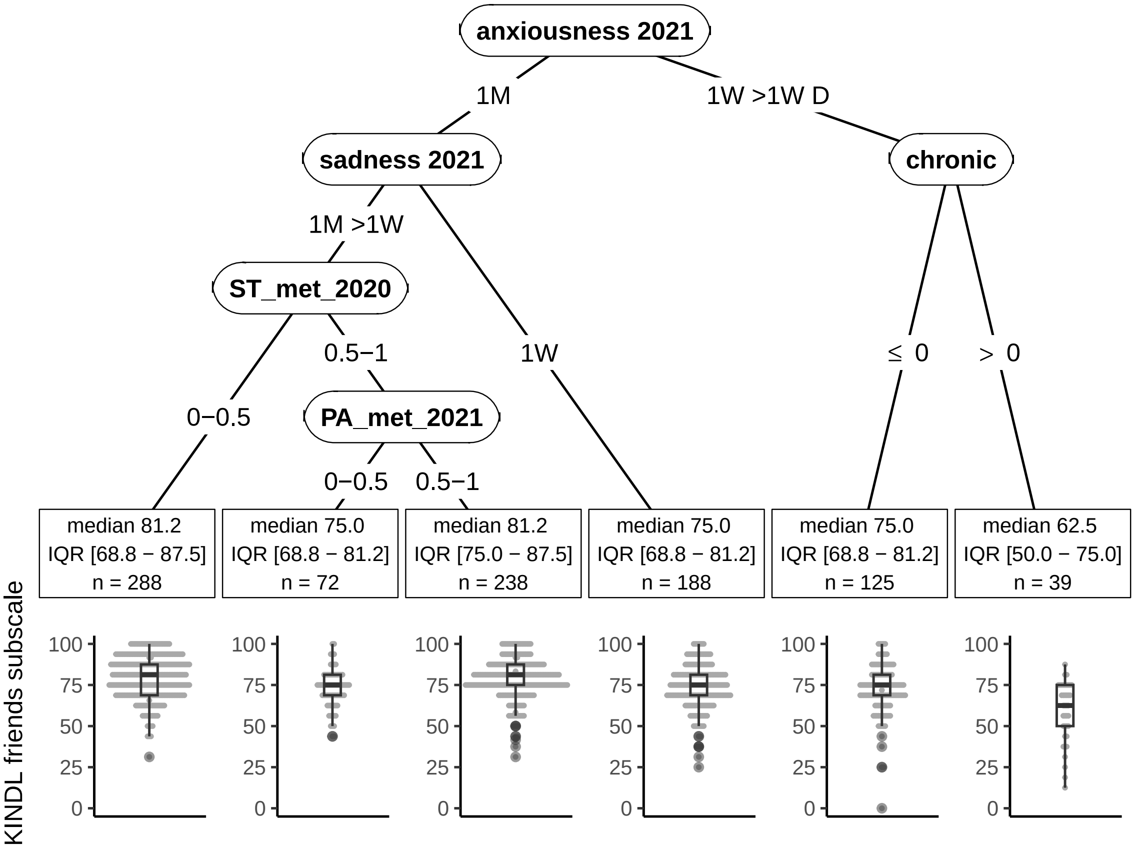

Figure S14: Model results for KINDL friends scale in secondary school children. See Section 3.2.

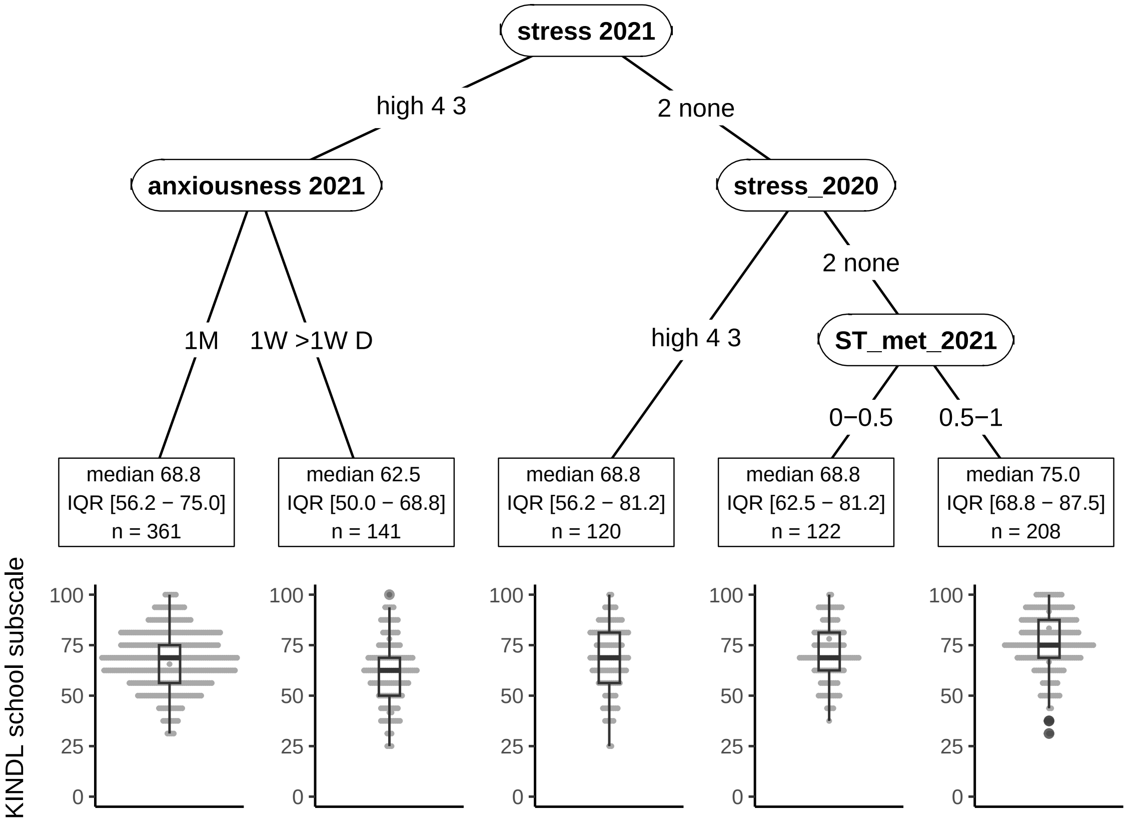

Figure S15: Model results for KINDL school scale in secondary school children. See Section 3.2.
